# Identifying bias in models that detect vocal fold paralysis from audio recordings using explainable machine learning and clinician ratings

**DOI:** 10.1101/2020.11.23.20235945

**Authors:** Daniel M. Low, Vishwanatha Rao, Gregory Randolph, Phillip C. Song, Satrajit S. Ghosh

## Abstract

**Introduction:** Detecting voice disorders from voice recordings could allow for frequent, remote, and low-cost screening before costly clinical visits and a more invasive laryngoscopy examination. Our goals were to detect unilateral vocal fold paralysis (UVFP) from voice recordings using machine learning, to identify which acoustic variables were important for prediction to increase trust, and to determine model performance relative to clinician performance.

**Methods:** Patients with confirmed UVFP through endoscopic examination (N=77) and controls with normal voices matched for age and sex (N=77) were included. Voice samples were elicited by reading the Rainbow Passage and sustaining phonation of the vowel “a”. Four machine learning models of differing complexity were used. SHapley Additive exPlanations (SHAP) was used to identify important features.

**Results:** The highest median bootstrapped ROC AUC score was 0.87 and beat clinician’s performance (range: 0.74 – 0.81) based on the recordings. Recording durations were different between UVFP recordings and controls due to how that data was originally processed when storing, which we can show can classify both groups. And counterintuitively, many UVFP recordings had higher intensity than controls, when UVFP patients tend to have weaker voices, revealing a dataset-specific bias which we mitigate in an additional analysis.

**Conclusion:** We demonstrate that recording biases in audio duration and intensity created dataset-specific differences between patients and controls, which models used to improve classification. Furthermore, clinician’s ratings provide further evidence that patients were over-projecting their voices and being recorded at a higher amplitude signal than controls. Interestingly, after matching audio duration and removing variables associated with intensity in order to mitigate the biases, the models were able to achieve a similar high performance. We provide a set of recommendations to avoid bias when building and evaluating machine learning models for screening in laryngology.

## INTRODUCTION

Voice recordings provide a rich source of information related to vocal tract physiology and human physical and mental health. Given advances in smartphones and wearables, these recordings can be made anytime and anywhere. Thus the search for disorder-specific acoustic biomarkers has been gaining momentum. Voice biomarkers have been reported for detecting Parkinson’s disease (1) as well as psychiatric disorders including depression, schizophrenia, and bipolar disorder (for a systematic review, see Low et al, 2020 (2)). Given our scientific understanding of the complexity of speech production, multiple acoustic features have been devised for use in machine learning models. In Figure 1, we describe a schematic of speech production and the process of extracting certain acoustic features from an audio signal (see also Quatieri, 2008 (3)), which is an important part of explaining how pathophysiology would affect acoustic features that are used in machine learning classifiers. Panel (A) depicts speech as the result of the neural coordination of three subsystems: the respiratory system (lungs), the laryngeal system (vocal folds), and the resonatory system of the vocal tract (pharynx, oral cavity, nasal cavity, articulators, and subglottal effects). Speech production requires air flow from the lungs to generate sound sources that are filtered by the vocal tract. Panel (B) captures the fact that environmental, microphone, and digital sampling characteristics (e.g., background noise, microphone gain, sampling rate) can affect acoustic features. Panel (C) shows the waveform of the audio signal, representing areas of compression (positive amplitude; higher air pressure) and rarefaction (negative amplitude; lower air pressure). Higher amplitudes can lead to higher perceived loudness. Prosodic features arise from changes over longer segments of time, which is perceived in the rhythm, stress, and intonation of speech. A segment of the waveform is shown in the right panel, indicating a periodic signal from the vocal folds. Panel (D) shows that for a given time window, a spectrum (right panel) can be obtained through a fast Fourier transform (FFT) which represents the magnitude of the frequencies in the signal with peaks (formants F1–F3) due to vocal tract filtering of the source signal produced by the vocal folds. The spectrogram (left panel) is a representation of the spectrum as it varies over time and can be obtained through a short-term Fourier transform (STFT). The approximate location of the F0 and first formants are displayed. Finally, (E) It is possible to separate source and filter components by computing the inverse FFT of the log of the magnitude of the spectrum, called the cepstrum (right panel). The peak in the cepstrum reflects the periodic glottal fold vibration while lower quefrency components reflect properties of the resonatory subsystem. For speech recognition, Mel filters are applied to the spectrum to better approximate human hearing. A conversion of the Mel-spectrum to a cepstrum using a Discrete Cosine Transform (DCT) generates mel-frequency cepstral coefficients (MFCCs). Similar to the cepstrum, lower MFCCs track vocal-tract filter information.

**Figure 1.**
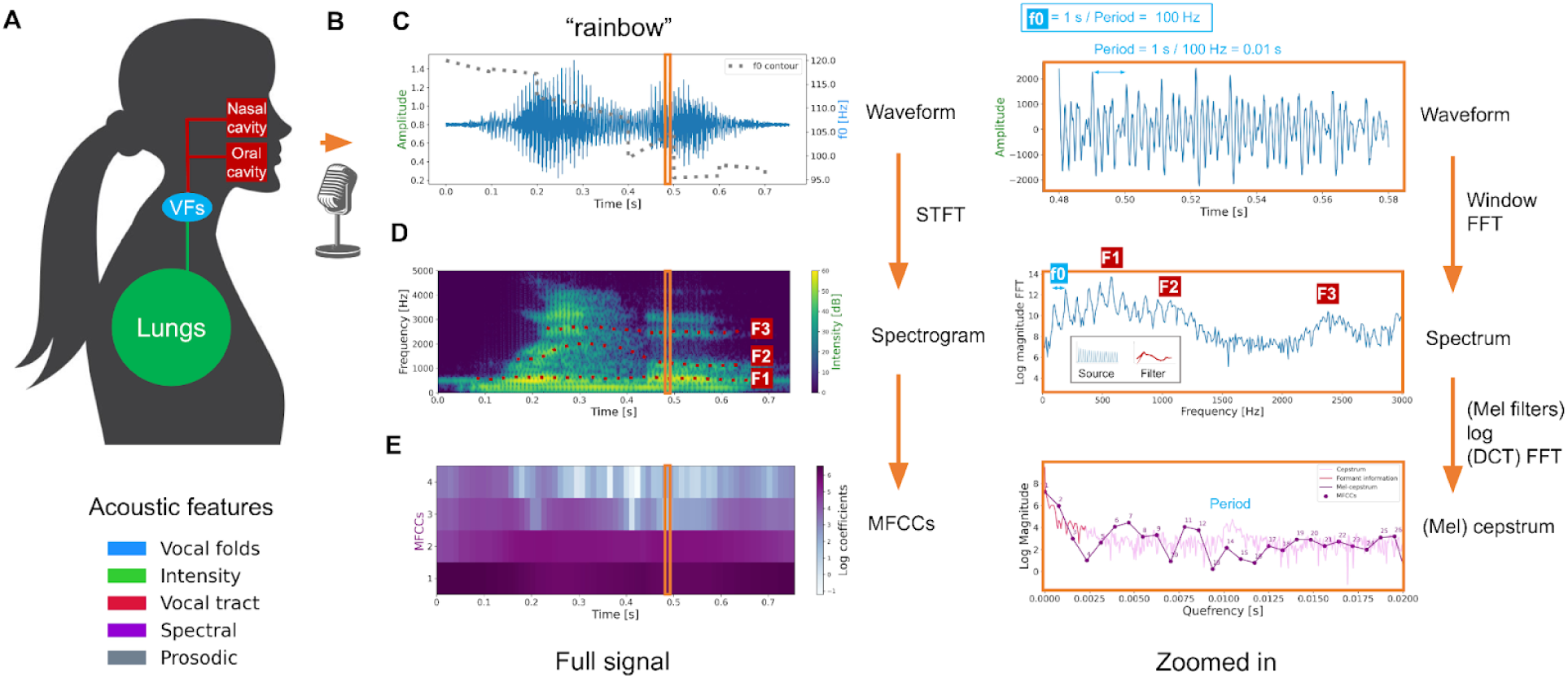
Schematic of speech production and the process of extracting certain acoustic features from an audio signal. (A) Speech production, (B) recording characteristics, (C) waveform of audio signal with fundamental frequency (f0), (D) spectrogram with formants F1-F3 and intensity, (E) mel-frequency cepstral coefficients (MFCCs). Full description in the main text.

Furthermore, while machine learning (ML) can be a powerful and successful approach for diagnostics, they are often treated as “black-boxes”. It can be difficult to determine how the model is making a decision, that is, how it is combining input features from a given patient to generate a prediction. This is particularly worrisome given ML algorithms can detect and associate unintended or clinically irrelevant relationships and introduce bias that may be difficult to anticipate. Explainable ML refers to a series of methods and quantitative analyses for uncovering and “explaining” the rationale behind the decision made by complex algorithms, which is particularly critical in the high-stake decisions of medicine to increase trust among clinicians and patients (4).

There are many challenges for applying acoustic analysis to detect specific disorders. Voice characteristics are highly varied and change over time. Laryngeal pathology, age, gender, size, weight, general state of health, smoking/vaping, and medications can impact vocal acoustic characteristics. Diseases in the larynx and phonatory system (i.e., larynx, resonating structures, lungs) and/or neurological system, will also affect voice. Compensatory production strategies and environmental conditions can also change the vocal signal. Furthermore, because hoarseness is such a frequent occurrence and specialty voice centers are rare, vocal fold disorders are often undiagnosed, under-reported, or misdiagnosed (5).

We chose vocal fold paralysis as the study cohort for several reasons. First, it is clinically important. UVFP can have detrimental effects on voice and quality of life with resultant morbidity related to respiration, swallowing and aspiration (6). Vocal fold paralysis may occur due to iatrogenic injury, malignancy, idiopathic, and neurological disease (7). Overall, surgical iatrogenic injury accounts for 46% of all UVFP in adults and thyroid and parathyroid surgeries are responsible for 32% of postsurgical UVFP (8). There is a significant need for a screening tool for the diagnosis and tracking of UVFP because of the high impact of this condition on productivity and quality of life. Screening could be done remotely and frequently, especially when surgical specialists and laryngeal exams are not readily accessible due to geographical, financial, and other barriers (9). Using an explainable ML model as a screening tool for UVFP can provide greater clarity as to who most needs laryngoscopy and provides insight in the key voice characteristics related to the pathophysiology (10–14). The costs associated with UVFP not only relate to patient morbidity and diminished quality of life but also to the economic burden placed on our healthcare system. Greater lengths of hospitalization and increased hospital costs have been associated with postsurgical VFP (15,16). Access to specialists for diagnosis is limited and early detection and management of UVFP appear to improve length of stay and surgical outcomes (17). Special consideration should be given to what the model can actually classify: a model that generalizes well in classifying UVFP from controls may not be able to screen for UVFP out of other voice disorders, but could be used to monitor UVFP patients remotely and affordably during treatment or detect risk for UVFP when it is the most likely cause such as dysphonia after thyroid surgery.

Furthermore, UVFP is an ideal model for demonstrating the explainability of ML. UVFP occurs when the mobility of a single vocal fold is impaired as a consequence of neurological injury and diagnosis is consistently verified through routine laryngoscopy; therefore, ground truth labels are available. Second, the clinical signs of UVFP are well-described. These characteristics include a weak, breathy voice quality, early vocal fatigue, reduced cough strength, and aspiration with thin liquids (18,19). Therefore, the acoustic differences between UVFP patients and healthy controls can be interpreted with regards to perceptual symptoms and a well-understood pathophysiology. In contrast, explaining important variables to predict a disorder which is hard to diagnose (e.g., has low inter-rater reliability) and has an unclear pathophysiology would ironically result in a poor explanation, because it would be puzzling how or even if the disorder could modulate the important acoustic variables. Of course, machine learning models can also offer novel explanations into a disorder by characterizing novel characteristics. However, if these models use high-dimensional feature vectors, they are more likely to overfit when using small datasets (20,21), which should lead to more skepticism of these novel explanations.

There have been several studies detecting unilateral vocal fold paralysis (UVFP) using machine learning (22–30); however, most have included the disorder among a set of voice disorders to be predicted. Limitations of these prior studies could be seen to fall into one of following types: not reporting the performance when classifying the subset of participants with UVFP out of the participants with dysphonia they were trying to detect; small sample sizes given most studies contained 10 participants with UVFP or fewer with one study containing 50 participants (31); a lack of algorithmic explanations: they either do not report on the relative importance of each acoustic variable; use input data such as a spectrogram in a black-box deep learning model which could make attempts at algorithmic explanations on images such as saliency maps more opaque than results from feature importance of handcrafted features; use a black-box model such as neural network without attempting to explain its predictions with deep learning explainability methods (32); use a single type of model which may pick up on certain types of patterns but miss others leading to incomplete conclusions on feature importance; use only a few features which may impede better predictive performance by not capturing certain relevant information; and/or not publicly share models or data to help test their generalizability to new data.

The objectives of our study were: to detect UVFP using ML; to evaluate the effectiveness of different models in differentiating the acoustic signals between patients with UVFP and patients with normal functioning vocal folds (i.e., controls); to explain which features are most important to the diagnostic models and examine the pathophysiological relevance; and to compare performance to human clinicians evaluating audio recordings. To achieve these objectives, we evaluated four different classes of machine learning algorithms to assess classification performance, obtained the minimal set of features necessary for detection, and identified the most important acoustic features for model construction after removing redundant features. Ultimately, we wanted to see if the most important features identified by the machine learning models matched clinically-known relevant acoustic changes.

## MATERIALS AND METHODS

This study was approved by the Institutional Review Board at Massachusetts Eye and Ear Infirmary and Partners Healthcare (IRB 2019002711).

### Participants and voice samples

Through retrospective chart analysis from 2009 to 2019, a total of 1043 patient charts were reviewed from a tertiary care laryngology practice who underwent endoscopic evaluation and voice testing. Of those, 53 patients with confirmed UVFP were identified. They had documented vocal fold paralysis by endoscopic examination and had undergone acoustic analysis as part of routine clinical care. Each patient had four acoustic recordings. These included three sustained vocalizations of the “a” vowel sound (ɑ in the International Phonetic Alphabet) and a reading of the introductory paragraph of the rainbow passage (33). The acoustic recordings were all taken in an acoustically shielded room. For each of these 53 patients, a board-certified otolaryngologist reviewed their clinical history, video laryngoscopy as well as their audio samples to confirm that they were correctly classified to have UVFP. Voice samples from an additional 24 patients were collected prospectively using a mobile software, OperaVOX^TM^ on an iPad, who were being treated for UVFP. These patients also had the same four acoustic recordings as the patients from retrospective chart review. This combination of data collection yielded a total of 77 UVFP patients for analysis, of which 48 had left UVFP and 29 right UVFP.

All of the patients were then matched with control samples from a database of patients without UVFP who had also undergone acoustic analysis. Each control was the same sex and had the same smoking status as the UVFP patient and within three years of age, and had documented laryngeal examinations that verified the absence of vocal fold mucosal pathology. The controls were excluded if they had established laryngeal surgery, vocal fold lesions, radiation, head and neck cancer, or neurological disease.

The controls had recorded the same four acoustic recordings as the retrospectively gathered UVFP group. A board-certified otolaryngologist confirmed that the voice recordings and video laryngoscopies of these controls matched normal expectancies. The reading samples were divided in thirds to match the amount of vowel production samples, resulting in 6 samples for most participants. Reading recordings were not available for three patients and three patient vowel samples were removed due to containing multiple vowel productions or a cough. The final dataset that was analyzed is described in Table 1. Reading+vowel refers to including all samples (i.e., ∼6 samples) from the same participant with the goal of either obtaining higher performance or discovering features that show variation in relation to diagnosis consistently across tasks. Mean (SD) audio lengths were 6.81s (5.47) for reading samples and 3.95s (1.00) for vowel samples. The audio samples were processed using OpenSmile with the eGeMAPS configuration file (article (34), source code (35)) which applies different summarization statistics to the time series depending on the feature resulting in 88 features per sample covering information related to the vocal folds (F0, jitter, shimmer), intensity (loudness, HNR), vocal tract (F1–3 frequency, bandwidth, amplitude), spectral balance (alpha ratio, Hammamberg index, spectral slope, MFCC 1–4, spectral flux), and prosody (voice and unvoiced segments, loudness peaks per second). See section “eGeMAPS features” in Sup. Mat. for full list.

**Table 1.** Sample sizes and demographic information.

|  | UVFP | Controls | Total |
| --- | --- | --- | --- |
| N | 77 | 77 | 154 |
| Mean age (SD) | 56.4 (18.7) | 56.6 (18.8) | 56.5 (18.7) |
| Sex (F/M) | 39/38 | 39/38 | 78/76 |
| Reading | 222 | 231 | 453 |
| Vowel | 227 | 231 | 458 |
| Reading+vowel (total) | 449 | 462 | 911 |

SD: standard deviation; F: female; M: male.

### Machine learning models of increasing complexity

With the goal of classifying voices recording into either UVFP or controls, we used four machine learning algorithms of increasing complexity from the *scikit-learn* (v0.21.3) using the *pydra-ml* (v0.3.1) toolbox (36) (default parameters were used unless otherwise specified). By complexity we mean models are more complex if they are harder to simulate, that is, harder to take the input data and model parameters and step through every calculation required to produce a prediction in a reasonable time which increases with the amount of parameters and interactions (37).

(1) Logistic Regression: a simple linear model that is constrained to use few features due to an L1 penalty making it the simplest model (“liblinear” solver was used which is ideal for smaller datasets).
(2) Stochastic Gradient Descent (SGD) Classifier: we used a log loss which implements a logistic regression; therefore, it is also a linear model but tends to use more features due to an elastic net penalty, making it slightly more complex (the max_iter parameter was set to 5000 and early_stopping was set to True).
(3) Random Forest: it is an algorithm that uses simpler decision trees (i.e., weak learners) on feature subsets “but then takes the majority of the votes of the decision trees’ predictions to create a stronger learner, making it harder to interpret which features are important across trees.
(4) Multi-Layer Perceptron: it is a neural network classifier which incorporates, in our case, 100 instances of perceptrons (artificial neurons), which are connected to each input feature through weights with a ReLU activation function to capture nonlinear relationships in the data. It is not possible to know exactly how the hundreds of internal weights interact to determine feature importance, making the model difficult to interpret directly from its parameters (the max_iter parameter was set to 1000; alpha or the L2 penalty parameter was set to 1).

To generate independent test and train data splits, a bootstrapped group shuffle split sampling scheme was used. Bootstrapping is more optimal than cross-validation on smaller datasets and provides a measure of uncertainty through a confidence interval (38). For each iteration of bootstrapping, a random selection of 20% of the participants, balanced between the two groups, was used to create a held-out test set. The remaining 80% of participants were used for training. This process was repeated 50 times, and the four classifiers were fitted and tested for each test/train split.. We used the default of 50 bootstrapping splits from pydra-ml to reduce computational time. Median ROC AUC stabilized to larger spit values at around 40 splits for logistic regression models across tasks (see Sup. Mat. Figure S1) while reducing runtime compared to larger split values. The Area Under the Receiver Operating Characteristic Curve (ROC AUC; perfect classification = 1; chance = 0.5) was computed to evaluate the performance of the models on each bootstrapping iteration, resulting in a distribution of 50 ROC AUC scores for each classifier. To ensure results were not due to choosing scikit-learn’s hyperparameter default settings, hyperparameter tuning was performed on the main models using all features and achieved similar performance to non-fine-tuned models (see Sup. Mat. Table S1). The focus of our study is identifying bias and not achieving –in our case– a small increment in performance; therefore, given the large number of models, analyses, and bootstrapping samples in our study which focuses on identifying bias, we chose default parameters given the small changes in performance we observed with hyperparameter tuning. Additionally, for each iteration, each classifier was trained with randomized patient/control labelings to generate a null distribution of ROC AUC scores (i.e., a permutation test). Each model’s performance was statistically compared to their null model’s distribution using an empirical p-value, a common and effective measure for evaluating classifier performance (see Definition 1 in (39)). The significance level was set to alpha = 0.05.

### Assessing feature importance

Kernel SHAP (SHapley Additive exPlanations) was used to determine which acoustic features were most important for each model to detect UVFP. This method is model agnostic in that it can take any trained target model (even “black box” neural networks) and compute feature importance (40). It does so by performing regression with L1 penalty between different sets of input features and a single prediction made by the target model. It then uses the coefficients of the additional regression model as a measure of feature importance for a single prediction. We took the average of the absolute SHAP values across all test predictions (positive and negative values are both important for classification). We then weighted the average values by the model’s median performance since an important feature for a bad model could be a less important feature for a good model and vice versa. Since we trained each model 50 times (i.e., one for each bootstrapping split), we computed the mean SHAP values across splits for each model. This pipeline (i.e., machine learning models, bootstrapping scheme, SHAP analysis) was done using *pydra-ml*.

### Reducing collinearity to do explainability analysis using Independence Factor

Highly correlated features (i.e., collinearity) can influence model generation and interpretation. Two models may obtain similar performance while using different features or placing different weights on the same features (i.e., underspecification (20,41)). This makes it difficult to compare algorithmic explanations across models. For instance, mean F1 frequency may be less important to a given model because the model uses mean F2 frequency which happens to capture very similar information in a particular dataset (i.e., has a high correlation), whereas a different model may use F1 instead of F2 or use both but assign less importance to each and still obtain the same performance. To enforce models to use the same features that capture very similar information and be able to compare feature importance across models, we kept a single feature out of the sets of features that share similar information above a given threshold.

We used a custom algorithm we call Independence Factor whereby for each feature in alphabetical (i.e., arbitrary) order, we removed features that show strong dependence above a given threshold. The step was repeated for remaining features. We use distance correlation from the Python *dcor* package (v0.4) because, unlike Pearson *r* or Spearman *rho*, it can capture non-monotonic relationships (42,43). We have included several examples of non-monotonic associations between variables in our dataset that would be captured better by dcor (see Sup. Mat. Figure S2). We used the following threshold values for the distance correlation [1.0, 0.9, 0.8, 0.7, 0.6, 0.5, 0.4, 0.3, 0.2] to compute the Independence Factor, which removed increasingly more features (i.e., 1.0 keeps all features and 0.2 removes features that have a distance correlation above 0.2). We chose the feature size which contains at least one model that scores within three percentage points of the performance using all features, with the goal of obtaining a more parsimonious model for subsequent explanation while maintaining high accuracy. Thus, removing redundant features makes the models easier to interpret for clinical relevance. To visualize the original redundancy across features, we computed clustermaps using *seaborn* package (v0.10.1) performing hierarchical clustering with the average-linkage method and Euclidean distance. This was performed on the pairwise distance correlation, computed separately on data from UVFP, controls, UVFP+controls and on reading, vowel, and reading+vowel.

### Performance using most important and least important features

Studies tend to report and describe the top N features out of M features, but it is not clear what performance the model would obtain when using only those top N features; perhaps it would perform substantially worse than the full model. We will report performance using only top 5 features as well as performance without top 5 features to provide a more realistic evaluation of their importance.

### Performance using audio duration

Figure 2 indicates clear differences in the distributions of audio recording duration between UVFP patients and controls. This is due to how recordings were processed and saved and not necessarily due to an intrinsic property of UVFP (e.g., slower speech), which reveals a bias that models can leverage but is not expected to generalize well under different audio processing procedures. Therefore, we examine whether audio duration alone could perform well in classification of UVFP. The mean (and standard deviation) for the audio duration for reading task is 3.5 s (0.00 s) for the controls and 10.25 s (6.17 s) for the UVFP patients and the audio duration for sustained vowel task is 4.11 s (0.07 s) for the controls and 3.74 s (1.3 s) for the UVFP patients.

**Figure 2.**
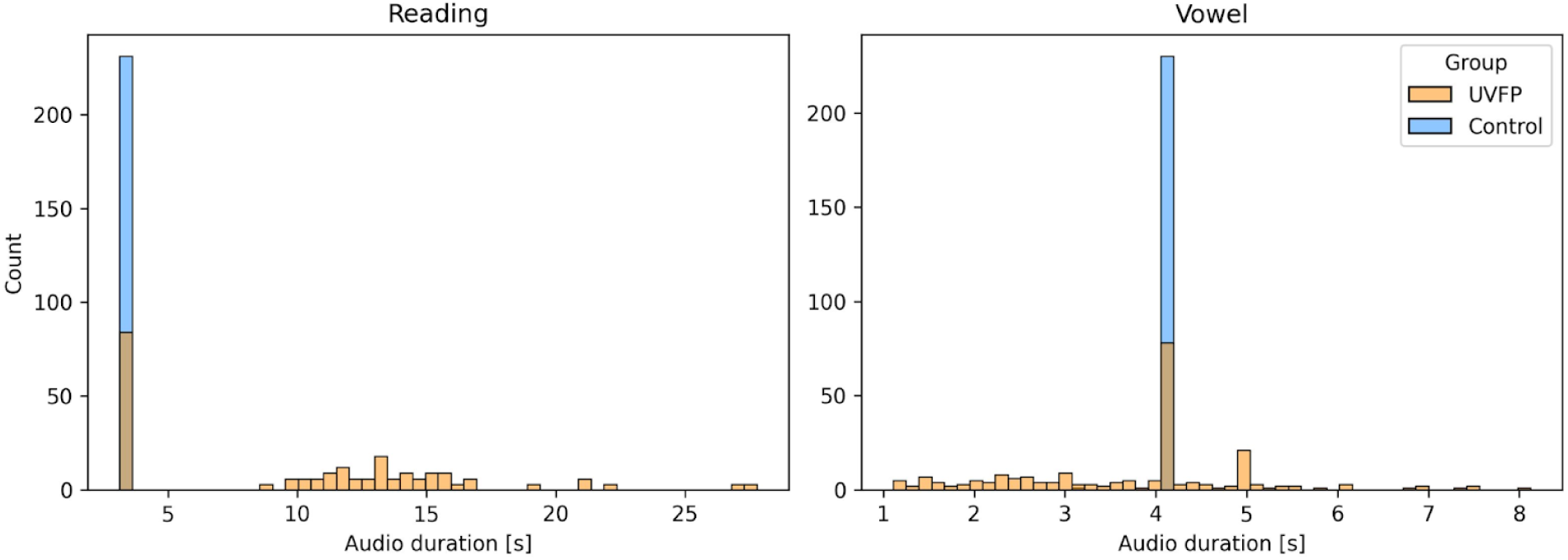
Distribution of audio duration for reading and vowel tasks split by group reveals a dataset bias. The mode of the audio durations for the controls is 3.5 s for reading samples and 4.11 s for vowel samples.

### Performance using cepstral peak prominence

To evaluate whether results are sensitive to choice of features, we use a different set of features derived from cepstral peak prominence (CPP) given it has been shown to be a good measure of breathiness and dysphonia (44,45). We match the summary statistics across the audio recording that eGeMAPS uses: CPP mean, CPP coefficient of variation (standard deviation normalized by the mean), CPP 20th percentile and CPP 80th percentile. We use our custom Python implementation which matches MatLab’s COVAREP output (46).

### Clinician ratings

In order to corroborate whether there are unintended recording differences between UVFP patients and controls that may lead to bias, one otorhinolaryngologist and two speech-language pathologists rated each audio recording of the reading task (one per participant, not split in three) for the following variables (and possible responses), in order: background noise (None, Some, High); UVFP (yes, no), CAPE-V severity (0 to 100), CAPE-V roughness (0 to 100), CAPE-V breathiness (0 to 100), CAPE-V strain (0 to 100), CAPE-V pitch (0 to 100), CAPE-V loudness (0 to 100; estimated loudness as if the rater were in the recording room), recording loudness (low, medium, high; loudness of the recording). Inter-rater agreement was assessed using intra-class correlation for all numerical variables and Light’s k for the binary presence of UVFP (47) using the R package *irr (v0.84.1)* (*48*). The entire reading task was provided instead of the task split in three to make assignment easier for clinicians. The reading task was chosen over the sustained vowel because we expected it to be easier for clinicians to detect UVFP.

## RESULTS

### Performance of models using acoustic features

In Table 2, we report performance for models using all features, models after removing redundant features, models using only top 5 features (to understand their unique role in performance), models using all 88 features without 5 features (to understand whether the top 5 features are necessary for high performance), models using audio duration length, and models using a different feature set based on CPP. Performance was found to be high across most models except CPP-based models. Some of the models just using audio duration length were able to achieve close to the highest performance, which reflects the expected effect of the difference in the dataset. Given dependent features provide similar information (see Supplementary Figures S1, S2, S3, S4, S5, S6, S7, S8, and S9) and distort feature importance analyses, we then tested performance after removing redundant features using the Independence Factor method previously described. Supplementary Figure S12 shows performance for different feature set sizes with increasing amounts of redundant features. From this analysis, we selected the feature-set size that resulted in best performance using the least amount of features for subsequent analyses: 39 features (reading), 13 (vowel), 19 (reading+vowel). After removing related features (i.e., reducing collinearity) from the original 88 features, similar performance was obtained (median ROC AUC = 0.84–0.87) using fewer features. Supplementary Materials “Feature selection” section describes an analysis of how this method compares to removing features across each train set (see Sup. Mat. Table S1).

**Table 2.** Model performance. Performance of models using either all 88 features, non-redundant features (39, 13, 19), top five most important features, all 88 features minus top 5 most important features using eGeMAPS features. We then compared this to using just audio duration as well as a different feature set based on CPP. Median ROC AUC score from 50 bootstrapping splits (90% confidence interval; median score of null model trained on permuted labels which should be at .50 if at chance). For full distributions of scores see Figure S11 in Supplementary Materials. Removing features is a post-hoc analysis because features were selected based on observing performance on the test sets, and therefore performance might be slightly overly optimistic and would need to be tested on an independent test set for further validation. MLP: Multi-Layer Perceptron; SGD: Stochastic Gradient Descent Classifier; CPP: Cepstral Peak Prominence.

|  | Features | LogisticRegression | MLP | RandomForest | SGDClassifier |
| --- | --- | --- | --- | --- | --- |
| Reading | 88 | .87 (.78–.93; .50) | .87 (.80–.93; .50) | .87 (.76–.91; .49) | .83 (.76–.89; .50) |
| Vowel | 88 | .84 (.77–.89; .50) | .86 (.79–.91; .50) | .86 (.79–.91; .51) | .80 (.72–.87; .50) |
| Reading+Vowel | 88 | .84 (.76–.91; .50) | .86 (.74–.92; .48) | .85 (.77–.92; .49) | .79 (.72–.86; .51) |
| Reading | 39 | .84 (.76–.92; .50) | .83 (.76–.91; .50) | .87 (.77–.91; .51) | .78 (.71–.86; .51) |
| Vowel | 13 | .80 (.70–.90; .50) | .81 (.74–.91; .50) | .84 (.75–.90; .52) | .74 (.58–.87; .51) |
| Reading+Vowel | 19 | .79 (.70–.84; .50) | .82 (.75–.88; .51) | .84 (.77–.91; .51) | .70 (.61–.77; .52) |
| Reading | Top 5 | .81 (.73–.89; .50) | .86 (.78–.92; .47) | .85 (.77–.90; .50) | .75 (.56–.87; .57) |
| Vowel | Top 5 | .78 (.67–.87; .50) | .82 (.74–.92; .53) | .81 (.72–.87; .50) | .72 (.57–.82; .49) |
| Reading+Vowel | Top 5 | .80 (.70–.86; .50) | .82 (.74–.88; .50) | .81 (.74–.89; .53) | .72 (.55–.83; .52) |
| Reading | 88 - Top 5 | .85 (.76–.92; .50) | .87 (.77–.92; .49) | .85 (.77–.90; .52) | .82 (.71–.89; .51) |
| Vowel | 88 - Top 5 | .84 (.75–.93; .50) | .86 (.72–.93; .51) | .84 (.74–.94; .52) | .80 (.70–.90; .48) |
| Reading+Vowel | 88 - Top 5 | .84 (.74–.89; .50) | .85 (.76–.91; .50) | .85 (.76–.91; .50) | .79 (.71–.87; .50) |
| Reading | Duration 1 | .81 (.73–.88; .50) | .81 (.73–.88; .50) | .85 (.77–.93; .50) | .76 (.50–.88; .50) |
| Vowel | Duration 1 | .70 (.61–.77; .50) | .80 (.70–.91; .51) | .86 (.76–.94; .52) | .50 (.31–.68; .51) |
| Reading+Vowel | Duration 1 | .70 (.64–.76; .50) | .76 (.67–.84; .50) | .86 (.73–.92; .50) | .64 (.45–.70; .50) |
| Reading | CPP 4 | .76 (.64–.84; .50) | .76 (.64–.84; .46) | .71 (.64–.78; .55) | .74 (.60–.84; .50) |
| Vowel | CPP 4 | .82 (.73–.90; .50) | .82 (.71–.90; .53) | .77 (.65–.85; .50) | .77 (.40–.86; .49) |
| Reading+Vowel | CPP 4 | .72 (.65–.80; .50) | .74 (.68–.84; .53) | .72 (.65–.78; .50) | .68 (.44–.78; .49) |

The bootstrapped ROC AUC distributions and permutation tests for the reduced (parsimonious) models using the non-redundant feature set are shown in Figure 3. Models distribution were all significantly different than their null distribution after correcting for multiple comparisons using a Benjamini-Hochberg procedure.

**Figure 3.**
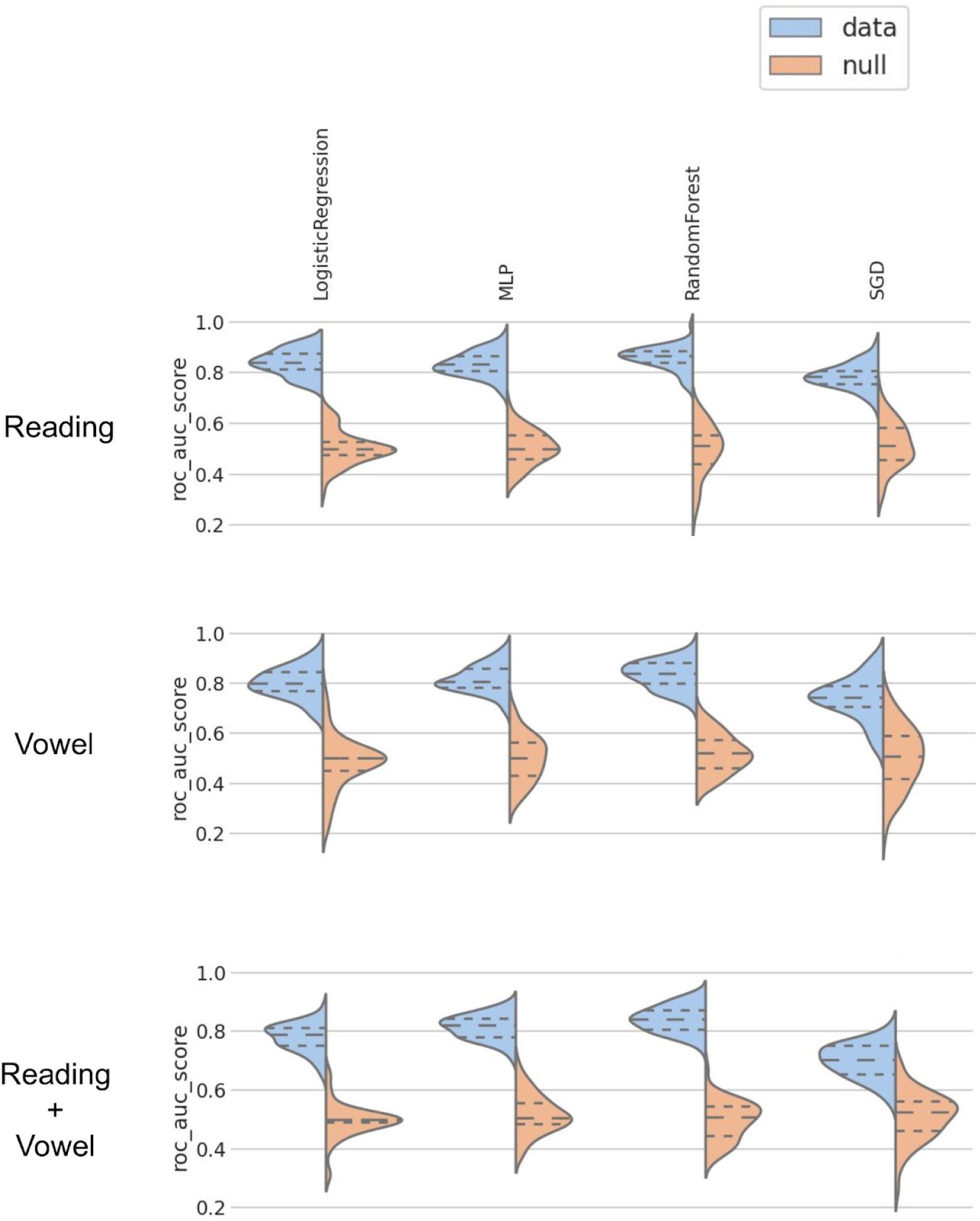
Model performance comparison using a permutation test using non-redundant features. Scores from models trained on true labels (blue) and trained on permuted labels (orange) over bootstrapping splits.

Given 24 UVFP patients were recorded with a different device, an iPad, we trained models without their samples to make sure these differences in recordings were not driving performance. There was a small drop in performance, which could be due to a bias (the full, original model using information of the recording device), but could also be due to removing training samples. The drop in performance is not large enough to suspect that differences in recording are driving the full original model’s performance (see Sup. Mat. Table S2, Table S3, and analysis in Supplementary section “Performance removing participants that used other recording system”).

### Assessing feature importance

Figure 4 reports feature importance using SHAP for all models. For the reading-based models, all models tend to use the same top 5 features except SGD, which also has the lowest performance. For further description of features and the chosen classification of features, see Eyben et al. (2015) (34) and Low et al. (2020) (2). When reviewing important features, it is key to note that any of the features with which it is codependent or associated could be a reasonable important feature (see clusters of redundant features in Supplementary Figures S3-S11). The variance on feature importance rank is evidence that models can use different feature information and still obtain similar high –although not perfect– performance. We further display the distribution of each top feature and its individual performance in Figure 5, which shows that no single feature is enough to dissociate groups with high performance. This figure also revealed the bias: the intensity-related feature equivalent sound level was counterintuitively higher for UVFP patients than controls. Figure 6 reports similarity between top 5 features and all original 88 eGeMAPS features. Features that have a high dcor or distance correlation (i.e., cluster) with top 5 features were not used in models to avoid redundancy, but still share similar information and can therefore be considered important features as well. Hierarchically-clustered heatmaps for other data types (vowel, reading, both) and groups (UVFP patients, controls, both) are displayed in Supplementary Figures S1, S2, S3, S4, S5, S6, S7, S8, and S9. Clustering tends to reflect pre-defined features types such as those reflecting patterns from vocal folds, intensity, vocal tract, spectral analyses, and prosody.

**Figure 4.**
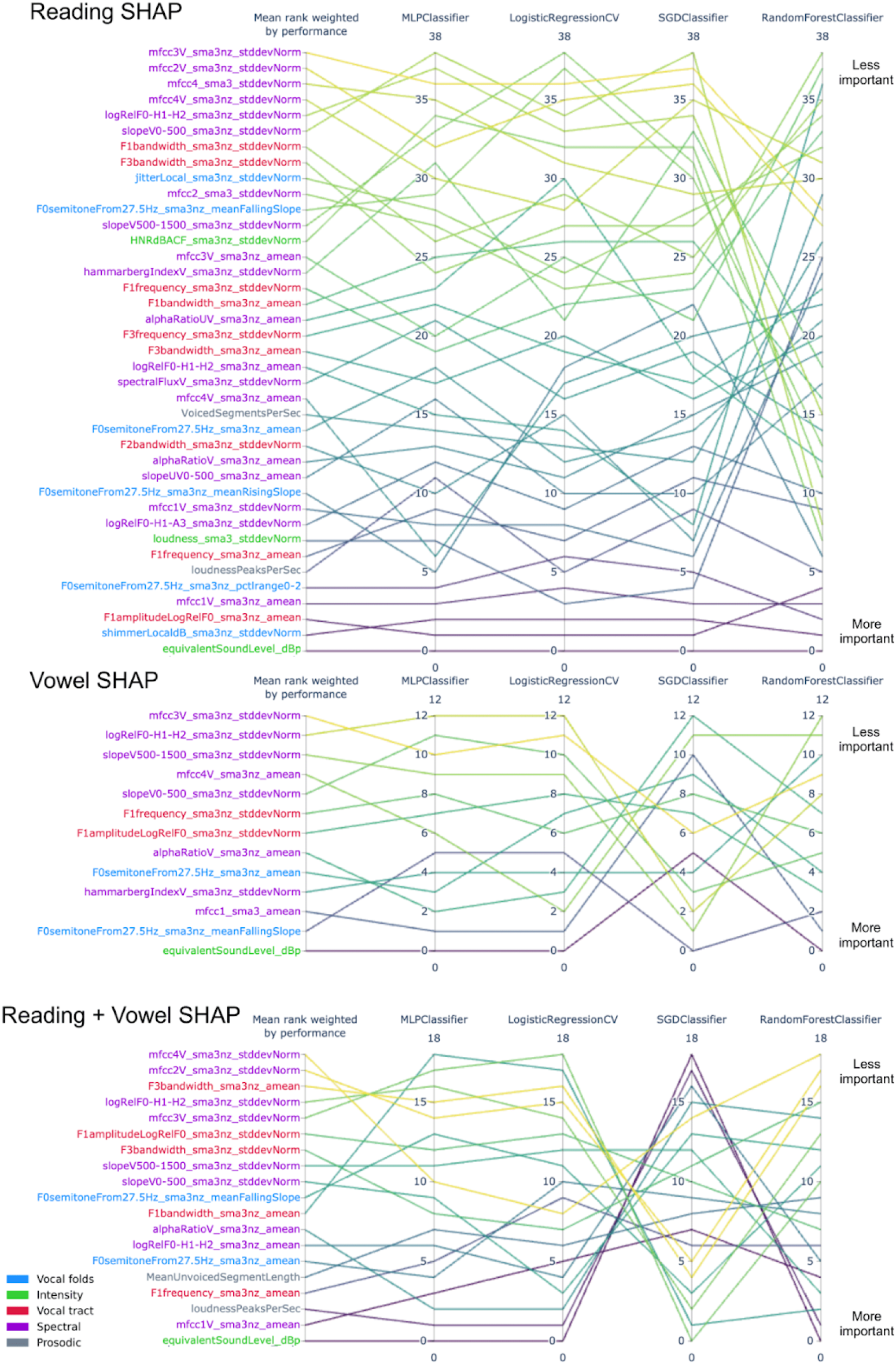
Feature importance parallel coordinate plot. Rank reads from bottom (most important) to top (least important). Mean rank is weighted by performance of each model to avoid a lower performing model biasing the mean rank.

**Figure 5.**
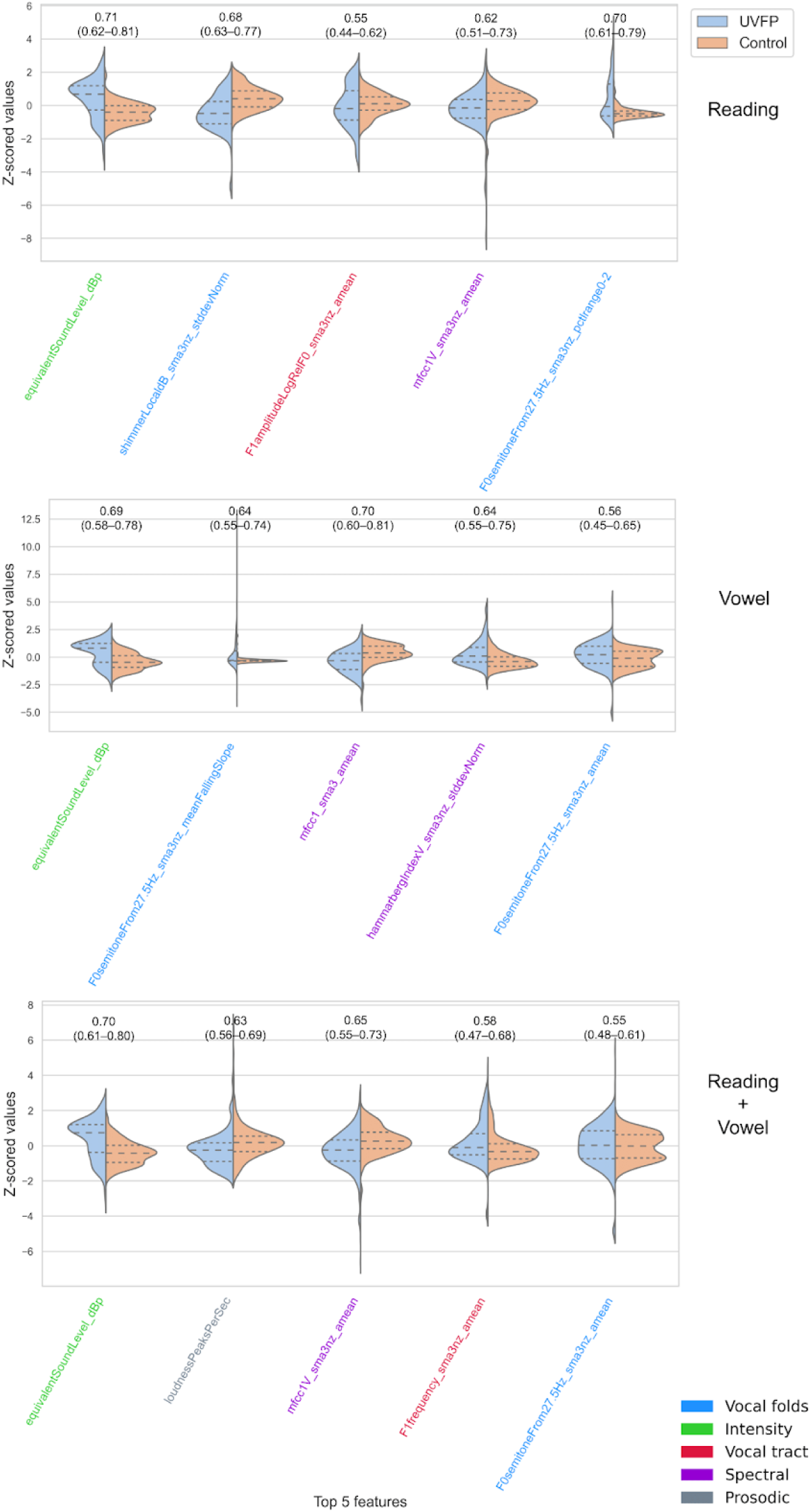
Distributions for top 5 features and corresponding performance for single features. Logistic Regression with L1 penalty was used. No single feature is enough to dissociate groups with high performance. Null models’ median performance was 0.5.

**Figure 6.**
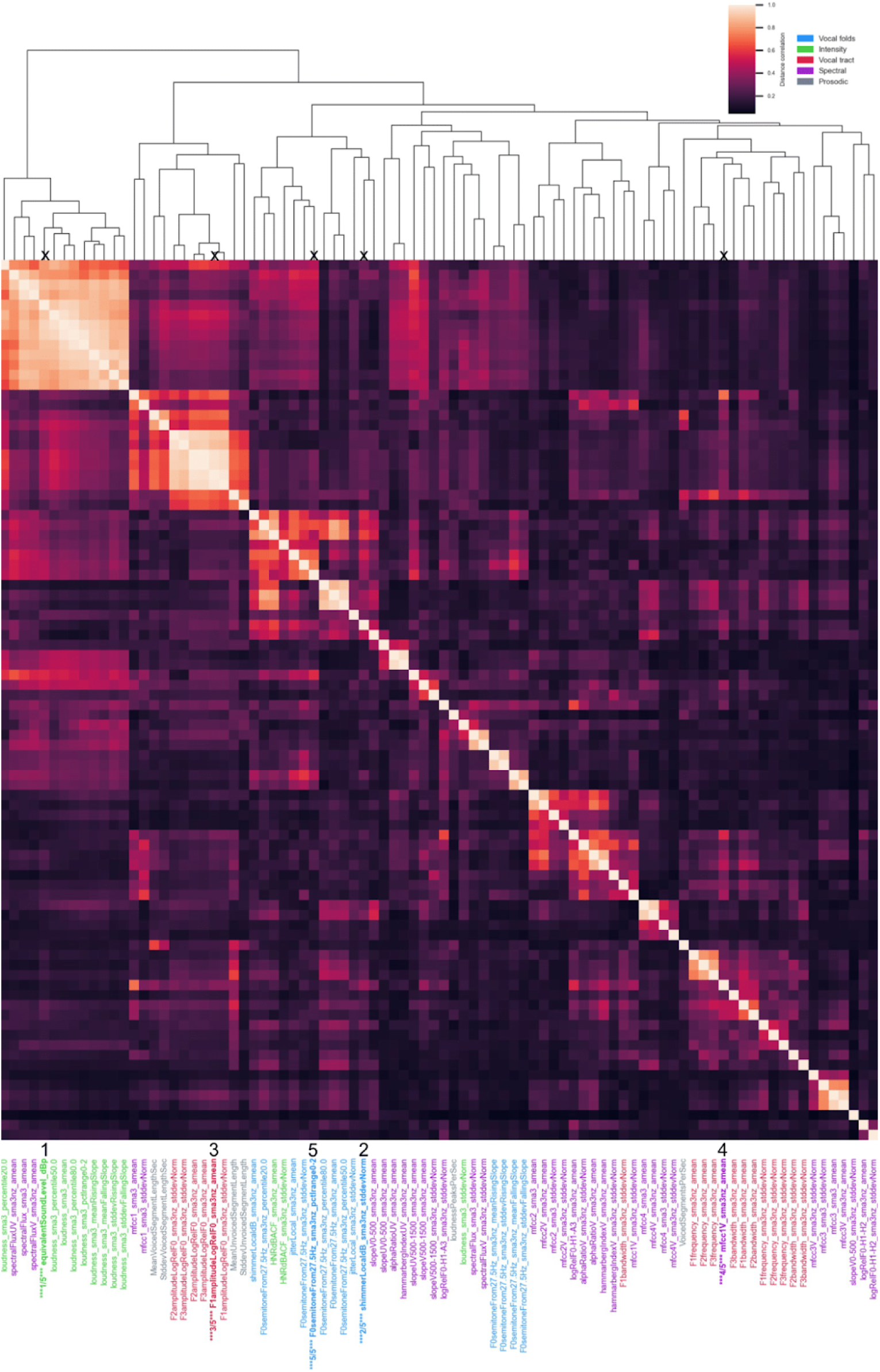
Feature redundancy with top 5 features highlighted. Top 5 features are highlighted in bold and their rank is displayed. Squares are clusters of redundant features. Computed with all participants on the reading task.

### Clinician ratings

The median ROC AUC for humans was 0.78 (min. = 0.74 to max. = 0.81) meaning the machine learning models performed better than the highest performing clinician on the limited available data, that is, the audio samples of the reading task. Interestingly, using the average clinician’s CAPE-V ratings within machine learning models was able to obtain a maximum median ROC AUC of 0.84 (0.72–0.92) with the Random Forest model (Table 3). Using clinicians’ perceptual ratings of background noise and recording loudness achieved a maximum median ROC AUC of 0.77 (.63– .87).

**Table 3.**
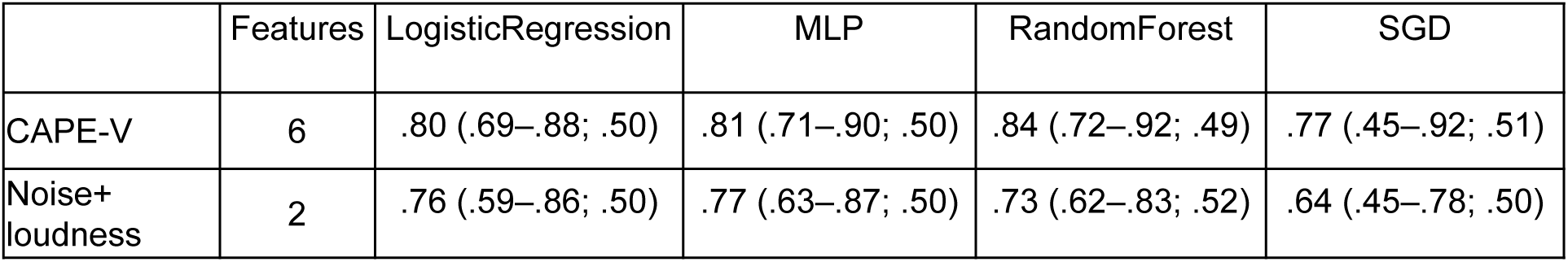
Performance using clinician ratings as variables for machine learning models. Median ROC AUC score from 50 bootstrapping splits (90% confidence interval; median score of null model trained on permuted labels which should be at .50 if at chance).

In Figures 6 and 7 we report the inter-rater reliability (Flight’s kappa and ICC) along with the distribution of the ratings. Common cutoffs for inter-rater agreement are poor for values less than .40, fair for values between .40 and .59, good for values between .60 and .74, and excellent for values between .75 and 1.0 (49). Background noise had poor reliability across rater, UVFP and recording loudness had fair reliability (see Figure 7) and CAPE-V-inspired ratings scored good to excellent except for pitch which was fair (see Figure 8).

**Figure 7.**
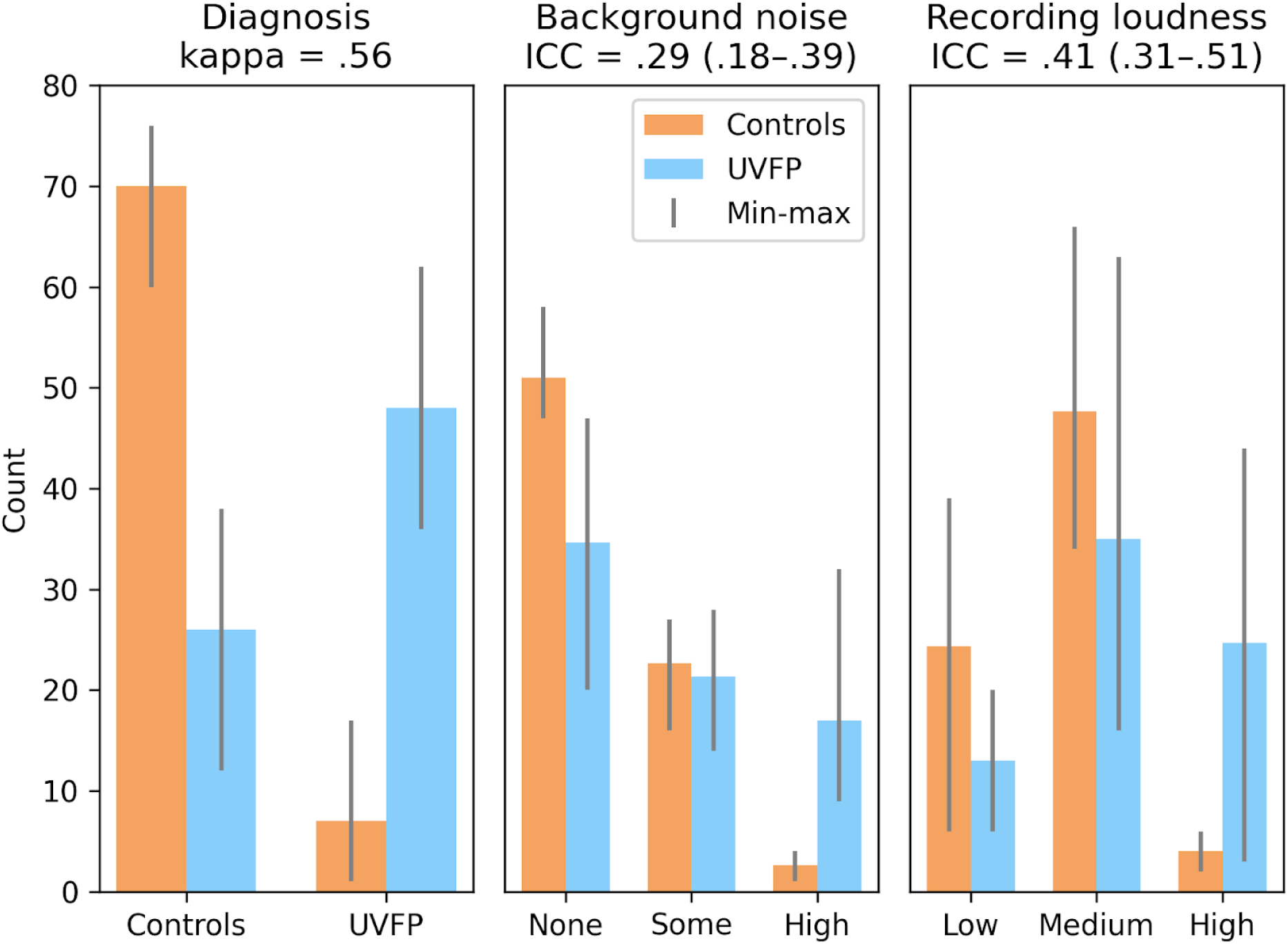
Descriptive statistics and inter-rater reliability of clinician ratings for unilateral vocal fold paralysis (UVFP), background noise, and recording loudness indicating likely bias. Controls and UVFP are ground truth diagnosis from the full clinical interview. Ratings are on brief reading samples. Bars indicate maximum and minimum count across the three raters. The disproportionate amount of UVFP samples rated as having high background noise and high loudness indicates likely bias, where the gain might have been raised for some UVFP patients and they may have phonated more intensely. kappa: Light’s kappa; ICC: intra-class correlation coefficient.

**Figure 8.**
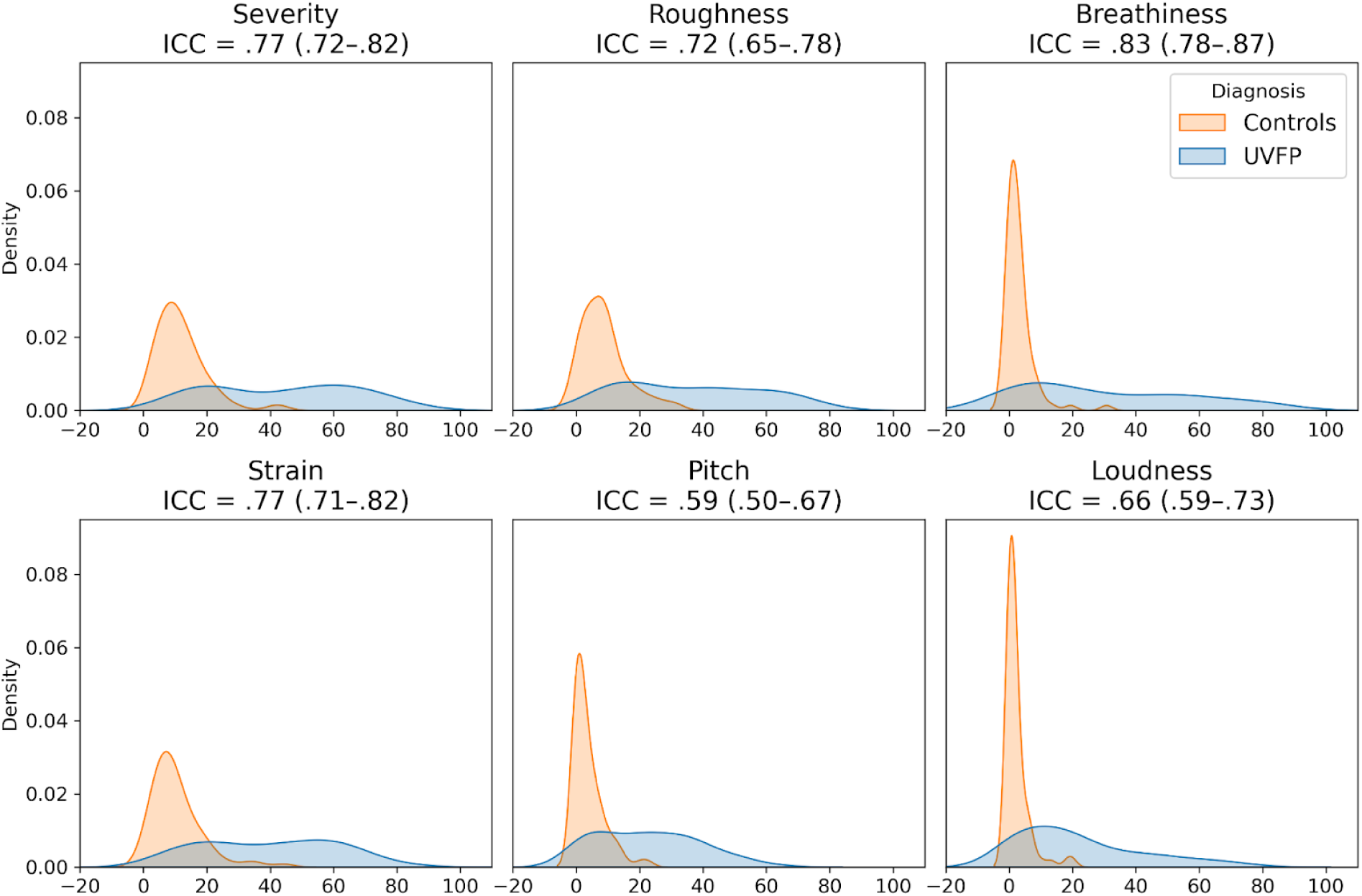
How clinicians rate the audio recordings of read speech: descriptive statistics and inter-rater reliability of average clinician ratings. The average across raters was taken for each recording. ICC: intra-class correlation coefficient.

### Bias mitigation: matching audio duration and removing features associated to intensity

We trimmed the longer UVFP samples so they were matched to control samples (all samples were the same duration), removing the audio duration difference. Vowel samples could not be matched by trimming as some UVFP samples were shorter and some were longer than control samples; therefore we demonstrate an attempt at bias mitigation only with reading samples. In Table 4, we show results on these samples after additionally removing all intensity features as well as variables that have a distance correlation (dcor) with any of them >= 0.3 and 0.4 based on the reading samples. Models have comparable performance to models with the original duration and intensity-related biases. See section “Biased features” and Table S4 in Sup. Mat. for a list of the 44 features associated with audio duration and the 14 intensity related features. For distance correlations between audio duration and features, see Sup. Mat. Table S6.

**Table 4.** Performance keeping features least associated with intensity features on samples of equal audio length after trimming.

|  | Features | LogisticRegression | MLP | RandomForest | SGD |
| --- | --- | --- | --- | --- | --- |
| dcor<0.4 | 44 | .88 (.80–.92; .50) | .87 (.81–.92; .47) | .87 (.78–.93; .45) | .83 (.76–.90; .48) |
| dcor<0.3 | 20 | .84 (.78–.89; .50) | .83 (.76–.9; .49) | .85 (.78–.91; .53) | .79 (.66–.87; .51) |

## Discussion

This study achieves high performance in detecting UVFP from healthy voices using a few seconds of audio recordings and surpassing clinician evaluations even after mitigating the biases we found in the dataset. As a result of performing the explainability analysis, we discovered a likely bias: intensity features were higher for UVFP patients than controls on average (Figure 5) when UVFP patients should have weaker voices. There are two likely causes. A first cause is that the software that had been used prompted users to speak louder if they had a weak voice in order to achieve an audible recording. A second cause was supported by clinicians’ ratings: clinicians rated UVFP patients as having louder recordings and more background noise than controls on average –when they should have similar levels–, which are proxies for microphone gain having been increased. This would have helped models improve performance using characteristics stemming from the recording idiosyncrasies instead of from pathophysiology. However, we removed features correlating with the clearly biased features and still achieved high performance.

Our study expands on prior studies which have used pre-existing commercial databases, smaller sample sizes, fewer features, and/or methods for model evaluation that can be biased in small datasets given the test sets may not be representative (for a discussion on bootstrapping for clinical datasets, see Figure 6 ^(2)^). Critically, we provide a roadmap for evaluating models more thoroughly including quantitatively explaining models and checking the robustness of the models to different choices of speech-eliciting tasks, algorithms, and feature sets. All of this should increase trust when no bias is found and when explanations are robust across models and make sense to experts. Such a model could fulfill several clinical needs: (1) postoperative screening for thyroid surgery-related UVFP since after thyroid surgery, UVFP is common, occurring in up to 5 to 10% of cases^27^. Furthermore, laryngoscopy is not readily available to all postoperative populations and symptomatic changes are notoriously variable. An ML-based screening could help identify patients needing further workup and treatment, and earlier diagnosis is essential to optimize long-term outcomes ^28,29^. (2) Monitoring voice during speech therapy and after surgical treatment for confirmed UVFP to measure when and if the patient’s voice is approximating a healthy voice. (3) Preoperative screening prior to surgeries that are at high risk for developing UVFP such as thyroid, head and neck, cardiac, thoracic, esophageal, and cervical spine operations.

In Table 5 we summarize several key recommendations to avoid bias when building and explaining machine learning tools for laryngology, although more could be added, and we expand upon how we dealt with some of these steps in the following sections.

**Table 5.** Recommendations to avoid bias for explainable machine learning models that use audio recordings for screening in laryngology.

| Recommendations | Description |
| --- | --- |
| Before data collection | - Pre-register hypotheses as to which variables should be important for predicting the target group to question effects that are not anticipated by theory (50) |
| During recording | - In a *controlled* recording setting: models could use any unintended differences between groups to improve classification (demonstrated in our study); therefore, it is important to make sure microphone gain, background noise, instructions are consistent across participants and reflect how recordings will be done once deployed.<br>- In a *remote* setting: it is desirable that models work on people's mobile devices outside the clinic. Since we cannot fully control the recording procedure, we should make sure there are no biases affecting one group more than another, test pilot instructions, and collect much more data to weaken the effect of individual recording idiosyncrasies.<br>- Perform pilot studies to do an initial quality control<br>- Collect representative samples so models generalize to different protected groups (e.g., ages, genders, races) or provide appropriate warnings (51).<br>- Providing instructions so participants do not overproject their voice and control recording procedure so a minimum loudness threshold is not needed (as demonstrated in our study) |
| Preprocessing and exploratory data analysis | - Quality control: remove non-natural outliers due to measurement errors, wrong data collection, or wrong data entry (e.g., fixing mislabeled files, unexpected silent recordings, recordings with extreme much background noise)(52)<br>- Avoid or be cautious with preprocessing steps that might reduce the properties associated with the disorder (e.g., denoising may remove breathiness information which may be useful for prediction).<br>- Observe distribution of variables between groups (e.g., audio duration) to make sure there are no differences that are not intrinsic to the disorder. Extra inspection of the data should be taken with retrospective studies where recording protocols were not controlled as in our study. |
| During training and evaluation | - Train multiple machine learning models of different complexity: two models may perform similarly but use input variables in different ways. If after training a model we only explain one of them, we might have biased conclusions of what variables characterize the disorder as we demonstrate.<br>- Avoid overfitting (i.e., finding patterns that do not generalize to new samples). Simple held-out test sets (e.g., of 20%) may not be representative of the population or the dataset, and therefore resampling methods (k-fold cross-validation, bootstrapping) are better. If performing hyperparameter tuning, nested resampling is needed to avoid overfitting (2). Avoid feature selection and dimensionality reduction using information from the test set/s. (38,53)<br>- Report performance on most and remaining important features as done in our study |
| During explainability analyses | - Choosing one of the variables that are highly dependent due to collinearity (e.g., that correlate above 0.8 Spearman rho or dcor above a threshold that does not reduce performance as we did in this study) or due to multicollinearity (remove variables if variance inflation factor > 5 or 10) (54); grouping correlated variables using leave-one-feature-out (LOFO); obtaining one variable from the correlated variables through dimensionality reduction (without using the test set which could lead to overfitting). |
|  | - Make conclusions from the features that are robustly important *across* models; here we take the average importance rank weighted by model performance.<br>- Evaluate potential bias: do important features match hypotheses? Do they dissociate groups in the expected direction? Do certain recording conditions perform better than others and were these done for only one group? Does the model work worse for certain races or age groups? Several metrics can evaluate this (e.g., see packages AIF360, fairlearn, and EqualityML).<br>- Use expert ratings to evaluate any potential sources of bias as done in our study.<br>- Understandability: are the explanations understandable for the engineer, the clinician, and/or the patient? (55) |
| If bias is detected | - Use bias mitigation strategies either during pre-processing (removing variables generating the bias along with variables correlated with these ones), training (adversarial debiasing, prejudice remover), or evaluation (equalized odds, reject option classification) (56). See packages AIF360, fairlearn, and EqualityML. |
| After deployment | - Continuous assessment: we need to review predictions and re-assess accuracy once deployed as new environments and populations could change performance (i.e., dataset shift (57)). |

### Explaining acoustic features relevant to detecting vocal fold paralysis

Objective acoustic measurement changes associated with vocal fold paralysis have been described and these changes include reduced loudness and maximum phonation time, higher perturbation measurements such as jitter and shimmer, and increased signal to noise ratio (19,58,59); however these were univariate models, and we have demonstrated that using single variables does not seem to provide high predictive performance. While other multivariate machine learning models have been used, these used few features and small or undefined samples and only report feature importance results for one model; therefore it is not clear whether the important features reported would hold using larger feature sets or how other models would perform. Using a much larger initial set of acoustic features for analysis, we demonstrate that several machine learning algorithms of increasing complexity (using more parameters) identify vocal fold paralysis from healthy voices. We also report that these models can use different features to achieve similar performance. Different models emphasize different features not simply because of its relevance to a disorder, but because of the mathematics associated with the model (e.g., containing different degrees of interaction effects, regularization, or propensity to underfitting or overfitting) (60). The variability of the ranking of features used by our individual models also illustrates the potential danger of using the single highest performing model, which is commonly seen in published literature.

Instead of simply reporting the important features from the highest performing model, we analyzed the models to find common features. The most important features across models were somewhat associated with intensity features (Sup. Mat. Table S5); therefore, even if not strongly associated with intensity features, they could be important due to a combination of intrinsic differences between UVFP and controls for which we provide hypotheses or because of how intensity influences them; a new unbiased dataset would be needed to confirm this. These top features were: intensity, especially equivalent sound pressure level which was redundant with multiple loudness features and seems to be due to some patients trying to use more breath for projection or being recorded with a higher microphone gain; Mel Frequency Cepstral Coefficients (especially the first coefficient, which captures spectral envelope or slope, which has be shown to be important for predicting UVFP ((29)); mean F0 semitones given F0 originates from vocal-fold oscillation, a vocal-fold paralysis is expected to alter F0, and has been shown to help predict pathological speech including UVFP (28);, mean F1 amplitude and frequency, influenced by how the vocal tract filters F0 and the shape of the glottal pulse which would be affected by UVFP voiced and unvoiced segments (prosodic and speech articulation features which may be altered due to changes in the periodicity of F0), and CPP features (which indicate voice quality degradations that could include more breathiness, a typical feature of UVFP (61)). Shimmer variability was important just for reading, and it captures variability in glottal pulses and pressure patterns which ultimately affect F0 and has been found to be significantly different between UVFP and a control group (62). When we removed the top 5 features from the full feature set, performance is practically equivalent to using 88 features, as expected, since there are features that are redundant with the top 5 features. Therefore, it is not that only these 5 specific features drive performance, but rather the information they contain, which in this dataset is also captured by other features as shown in Figure 6.

These acoustic features would corroborate our clinical understanding of glottal incompetence from UVFP and with common patient complaints of reduced loudness, vocal instability, hoarseness, and rough voice; however, they could also be important due to their associations with intensity features. Uncovering and understanding the basic mechanisms and features that models use to generate predictions and outcomes are important as these tools become part of the clinical decision making process.

### Identifying and addressing bias

Equivalent Sound Level was higher in UVFP patients than controls. This is counter-intuitive because UVFP patients are known to have softer voices as already described; however, clinicians rated most UVFP samples as being louder than controls. The bias discovered was likely due to increasing the gain on the microphone for some UVFP patients, which would explain the increased background noise in UVFP patients’ recordings. A second source of bias may have occurred from requesting UVFP patients to speak louder in order to meet the minimum intensity threshold on the recording softwares Computerized Speech Lab™ and OperaVOX, or patients could have tried this on their own knowing they were being recorded. This behavioral compensation is likely to occur in biomarker research when the participant has a soft voice, especially in retrospective studies like ours where the study goal is not known at the time of recording or when certain software properties lead individuals with weak voices to speak louder. Even though the current models perform better than the clinicians, a systematic comparison would require more clinician and model assessments across datasets. It is likely a model trained on a single dataset might learn intrinsic characteristics of that dataset that do not generalize as well as clinical expertise might.

Having said this, this line of research would help us understand the extent to which UVFP detection is generalizable from acoustic data alone. Finding an objective measure of hoarseness is important given a “normal voice” is a fundamentally subjective classification that is not well defined (63,64) and varies with training (65,66), which may result in low reliability of evaluation of disordered voices among clinical rating scales (67).

As a post hoc analysis, we address bias by trying to mitigate its effect: we removed variables associated with intensity variables on samples matched on audio duration. After removing these features, the models were able to obtain similar performance using a very different set of features. It is possible that these remaining features better reflect pathophysiology or that the features extracted are still influenced by intensity, but further studies should address their generalizability or their relation to intensity variation.

### Evaluating the sensitivity to tasks, model complexity, and features used

In addition to getting a better understanding of features, we explored performance in the context of different vocal tasks. Participants carried out two different tasks to elicit voice, *reading*, which captures more complex speech dynamics, and *sustaining vowels*, which is a simpler measure of vocalization and the respiratory subsystem. Overall, these dynamics from the speech task may have improved model performance as was observed.

Comparing simpler and more complex models is important because simpler models such as Logistic Regression could be preferred because they tend to generalize better given they are less at risk for overfitting the training set and they are more interpretable and thus biases can be assessed more directly (68).

By removing redundant features, we can concentrate on finding the most useful features for further analysis. Performance decreased only slightly while we made models more parsimonious and explainable. This approach is key given the curse of dimensionality in machine learning that may make models unnecessarily complex and harder to generalize (20).

Often studies will report the top N features but not how predictive they are in isolation. In our study we ran models on the top 5 features together (Table 2). The lower performance of these top 5 features relative to a richer feature set helps demonstrate that model performance is dependent on interactions across multiple additional features (with the exception of samples from the reading task which obtained an AUC of 0.86 using just the 5 features). We also ran models without top 5 features to demonstrate that leaving features that are redundant with these top features results in almost equivalent high performance to using all 88 features since the redundant features share information. Furthermore, when training models on the individual features from within these top 5 one at a time, the performance was reduced considerably with scores from 0.55 to 0.71. This indicates the need for these models to combine multiple features to achieve high performance and any model evaluation should not focus on only the common or top features without testing their predictive performance.

### Limitations and future directions

We cannot determine how the bias will affect the model’s performance on future samples, but it will likely underperform in samples where length was not different between groups, where gain cannot be changed, and where participants are instructed to not overproject their voice; however, it is possible the model could underperform for other reasons including dataset shift (e.g., the distribution of voice characteristics or demographics is different in a new sample).

The classification using just duration itself varied across models and clinicians who listened to the reading passage in its entirety did not achieve as good a classification as the best performing models. Duration itself was not included as a feature in the eGeMaps-based models and has a complex effect on both machines and humans. For example, duration could have affected eGeMAPS features (e.g, introduce more variability to the functionals that are computed over sliding time windows) and duration of vowels varied extensively across the UVFP group thus cannot itself be tied to underlying pathophysiology. Therefore, important future work should analyze how duration may affect these features, should address the intrinsic variability in durations of UVFP patients in responding to speech items, and should incorporate models of production that include a consideration of respiratory capabilities, articulation changes, and vocal fold pathophysiology.

It is not clear whether these models could detect UVFP from other voice disorders or just healthier voices; however, a model that generalizes well in classifying UVFP from controls could be used to monitor UVFP patients remotely and affordably during treatment or detect risk for UVFP when it is the most likely cause (e.g., dysphonia after thyroid surgery). Larger sample sizes with curated examinations can help increase diverse representation across voice quality and thereby potentially reduce bias in classifier performance. We did not analyze potential racial bias given this data was not extracted from the chart review. Our choice of a standardized feature set worked well in this setting, but may fail to work for differential voice disorder diagnosis or when generalizing to larger datasets, which may bring in additional sources of variance unaccounted for in this dataset. With the availability of more data, additional features could be extracted that better capture changes in coordination (e.g., XCORR (69)).

Furthermore, while our feature importance evaluation method, SHAP, shows a certain amount of robustness across models, alternative model-agnostic feature-importance methods (e.g., LOFO, permutation importance) as well as model-specific methods (coefficient values for linear models, mean decrease in impurity for Random Forest) could be compared. Model understandability –how easily are the explanations understood by a speech scientist or a clinician– could be assessed rigorously (55).

Finally, debiasing the models by removing features correlated with the biased ones was attempted although it is not clear how exactly intensity may influence certain features; we assume if intensity is influencing a variable, it generally should create some considerable association which we discarded using dcor. Therefore, the effect of the bias can be assessed by testing the model’s generalizability to new unbiased datasets. Therefore, we are not promoting our final debiased models as completely unbiased or ready to use, it is possible our debiasing strategies are only partially effective, additional biases remain, and/or additional ways of debiasing have not been considered.

We tested how well a model using only the top 5 features performed independently of the model with all features; it is possible to also test how well the incremental set of top features performs (1st, 1st and 2nd, 1st–3rd, etc.), which would be useful in order to compare different models’ performance as a function of which features are being used.

## Conclusion

Using one of the largest UVFP datasets to date, our study demonstrates the importance of checking for biases using explainable machine learning and clinician perceptual ratings. In order to first explain models, we tackle collinearity (i.e., redundant or highly correlated independent variables), which biases feature importance, using a custom method called Independence Factor that selects one out of a set of associated features without losing predictive performance. We then compare how results change across different speech-eliciting tasks, training algorithms, features, features set sizes, and highest and lowest performing features to better understand the process that models use to predict vocal changes associated with laryngeal disease, since analyzing a single model will result in a biased view of how predictions are achieved. During this process, we discovered there was a difference in audio duration between groups clearly not related to intrinsic differences in UVFP speech rate, but in cropping all control recordings to a certain length during audio storage. We also discovered that sound equivalent level was counterintuitively higher in UVFP patients, a likely bias resulting from the weak or underprojected voice that characterizes many UVFP patients: patients were prompted by the recording software to speak louder and the microphone gain was likely raised selectively for these patients with weaker voices, possibly generating higher background noise which was detected through clinician’s ratings; therefore the models picked up on the acoustic correlates of this increased intensity, which would impede generalization under different recording procedures and natural audio durations. This is more likely to occur in laryngology datasets when patients have a softer voice.

Interestingly, we found that matching audio duration between groups and removing all variables that were clearly related to intensity (e.g., bias mitigation) resulted in similar high performance. In this case, the model may be using information more related to pathophysiology, which would need to be further confirmed by future unbiased samples. Machine learning models tended to surpass clinician’s evaluation of the same audio recordings. Interestingly, using clinician’s voice quality ratings on the recordings in machine learning models performed better than their binary evaluation on whether recordings contained a sample of UVFP voice or not.

We hope to promote moving beyond using a single model and only reporting top features to a better explanation of how these models work as well as being able to understand variance across modeling and evaluation choices. We believe these are all aspects of machine learning that clinicians need to understand prior to using such applications.

With these considerations along with the recommendations we make, machine learning applications could aid in laryngology screening, allowing for the potential development of in-home screening assessments and continuous pre- and post-treatment monitoring.

## Data Availability

All data and code are available through Github (https://github.com/danielmlow/vfp) and Zenodo (https://doi.org/10.5281/zenodo.5009208) including a tutorial to test our models on your own data (https://github.com/danielmlow/vfp/blob/main/vfp_detector.ipynb).

https://github.com/danielmlow/vfp

## Acknowledgments

We would like to thank Cody Sullivan and Carolyn Hsu for their help in rating the audio samples and thank Daryush Mehta, Robert Hillman, and John Guttag for their feedback on an earlier version of this study. DML was supported by a National Institute on Deafness and Other Communication Disorders T32 training grant [5T32DC000038-28], a RallyPoint Fellowship, and an Amelia Peabody Professional Development Award. The work was supported by a gift to the McGovern Institute for Brain Research at MIT. SSG was partially supported by National Institutes of Health grants for the development of pydra-ml [R01 EB020740], for reproducible practices [P41 EB019936], and the Bridge2AI voice data generation project [1OT2OD032720-01]. The authors declare that there is no conflict of interest.

## Author Contributions

Daniel M. Low: Data curation, Methodology, Formal analysis, Software, Writing - Original Draft; Vishwanatha Rao: Data Curation, Formal analysis, Writing - Original Draft; Gregory Randolph: Writing - Review & Editing; Philip C. Song: Conceptualization, Methodology, Writing - Original Draft, Supervision, Data curation; Satrajit S. Ghosh: Conceptualization, Methodology, Writing - Original Draft, Supervision, Software

## Notes

### Competing Interest Statement

The authors have declared no competing interest.

### Summary of Updates

Figure 1 and 3 updated.

