## Supplementary Materials for "Identifying bias in models that detect vocal fold paralysis from audio recordings using explainable machine learning and clinician ratings"

### METHODS

#### List of eGeMAPs features

**Here is a list of features used from eGeMAPS (see main manuscript for citation and source code):**
F0semitoneFrom27.5Hz_sma3nz_amean,
F0semitoneFrom27.5Hz_sma3nz_meanFallingSlope,
F0semitoneFrom27.5Hz_sma3nz_meanRisingSlope,
F0semitoneFrom27.5Hz_sma3nz_pctlrange0-2,
F0semitoneFrom27.5Hz_sma3nz_percentile20.0,
F0semitoneFrom27.5Hz_sma3nz_percentile50.0,
F0semitoneFrom27.5Hz_sma3nz_percentile80.0,
F0semitoneFrom27.5Hz_sma3nz_stddevFallingSlope,
F0semitoneFrom27.5Hz_sma3nz_stddevNorm,
F0semitoneFrom27.5Hz_sma3nz_stddevRisingSlope, F1amplitudeLogRelF0_sma3nz_amean, F1amplitudeLogRelF0_sma3nz_stddevNorm, F1bandwidth_sma3nz_amean, F1bandwidth_sma3nz_stddevNorm, F1frequency_sma3nz_amean, F1frequency_sma3nz_stddevNorm, F2amplitudeLogRelF0_sma3nz_amean, F2amplitudeLogRelF0_sma3nz_stddevNorm, F2bandwidth_sma3nz_amean, F2bandwidth_sma3nz_stddevNorm, F2frequency_sma3nz_amean, F2frequency_sma3nz_stddevNorm, F3amplitudeLogRelF0_sma3nz_amean, F3amplitudeLogRelF0_sma3nz_stddevNorm, F3bandwidth_sma3nz_amean, F3bandwidth_sma3nz_stddevNorm, F3frequency_sma3nz_amean, F3frequency_sma3nz_stddevNorm, HNRdBACF_sma3nz_amean, HNRdBACF_sma3nz_stddevNorm, MeanUnvoicedSegmentLength, MeanVoicedSegmentLengthSec, StddevUnvoicedSegmentLength, StddevVoicedSegmentLengthSec, VoicedSegmentsPerSec, alphaRatioUV_sma3nz_amean, alphaRatioV_sma3nz_amean, alphaRatioV_sma3nz_stddevNorm, equivalentSoundLevel_dBp, hammarbergIndexUV_sma3nz_amean, hammarbergIndexV_sma3nz_amean, hammarbergIndexV_sma3nz_stddevNorm, jitterLocal_sma3nz_amean, jitterLocal_sma3nz_stddevNorm, logRelF0-H1-A3_sma3nz_amean, logRelF0-H1-A3_sma3nz_stddevNorm, logRelF0-H1-H2_sma3nz_amean, logRelF0-H1-H2_sma3nz_stddevNorm, loudnessPeaksPerSec, loudness_sma3_amean, loudness_sma3_meanFallingSlope, loudness_sma3_meanRisingSlope, loudness_sma3_pctlrange0-2, loudness_sma3_percentile20.0, loudness_sma3_percentile50.0, loudness_sma3_percentile80.0, loudness_sma3_stddevFallingSlope, loudness_sma3_stddevNorm, loudness_sma3_stddevRisingSlope, mfcc1V_sma3nz_amean, mfcc1V_sma3nz_stddevNorm, mfcc1_sma3_amean, mfcc1_sma3_stddevNorm, mfcc2V_sma3nz_amean, mfcc2V_sma3nz_stddevNorm, mfcc2_sma3_amean, mfcc2_sma3_stddevNorm, mfcc3V_sma3nz_amean, mfcc3V_sma3nz_stddevNorm, mfcc3_sma3_amean, mfcc3_sma3_stddevNorm, mfcc4V_sma3nz_amean,

mfcc4V_sma3nz_stddevNorm, mfcc4_sma3_amean, mfcc4_sma3_stddevNorm, shimmerLocaldB_sma3nz_amean, shimmerLocaldB_sma3nz_stddevNorm, slopeUV0-500_sma3nz_amean, slopeUV500-1500_sma3nz_amean, slopeV0-500_sma3nz_amean, slopeV0-500_sma3nz_stddevNorm, slopeV500-1500_sma3nz_amean, slopeV500-1500_sma3nz_stddevNorm, spectralFluxUV_sma3nz_amean, spectralFluxV_sma3nz_amean, spectralFluxV_sma3nz_stddevNorm, spectralFlux_sma3_amean, spectralFlux_sma3_stddevNorm.

#### Choosing amount of bootstrapping splits

We chose the default of 50 bootstrapping splits from pydra-ml. Additional testing shows median ROC AUC stabilized to larger spit values at around 40 splits for logistic regression models across tasks for using 88 features (see Figure S1).

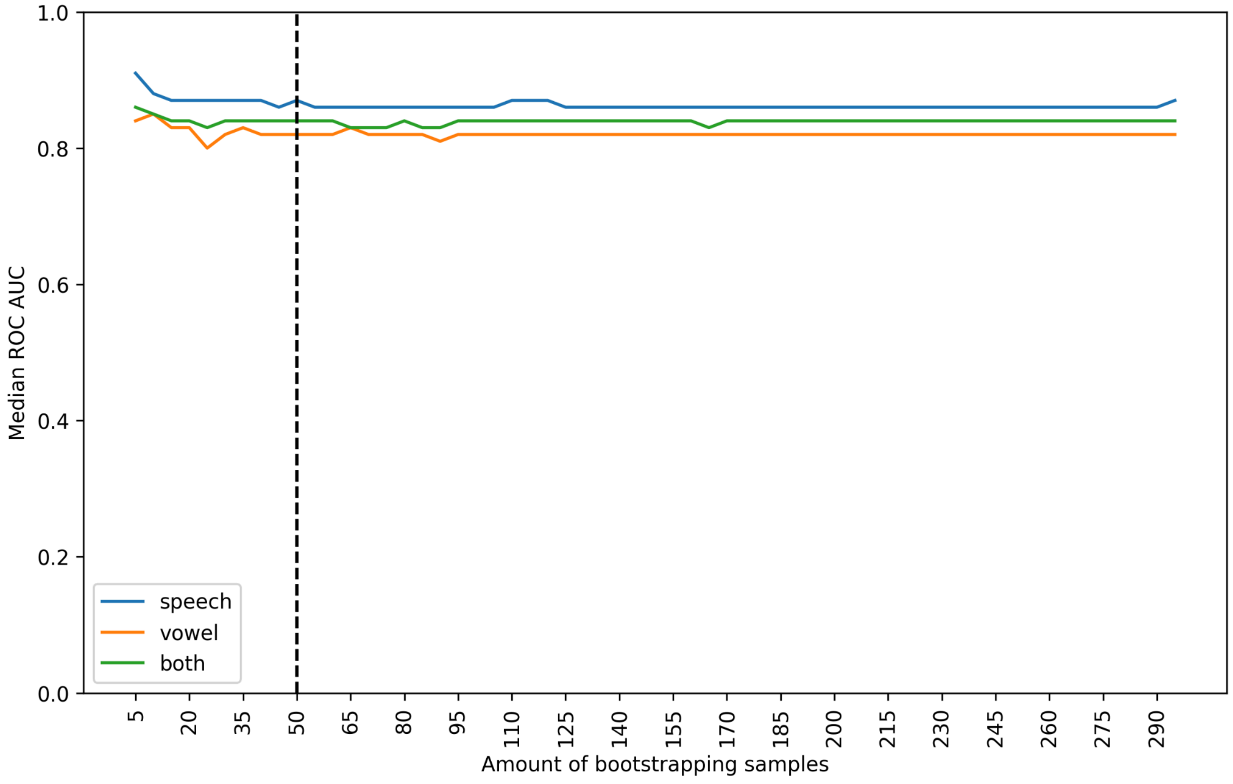


**Figure S1.** Controls, reading+vowel tasks: Visualization of features with shared information using pairwise distance correlation across the 88 features. Squares are clusters of redundant features.

#### Non-monotonic association captured better by distance correlation

See Figure S2 for non-monotonic associations. We highlight the trend using generalized additive models from R's ggplot2 geom_smooth() function.

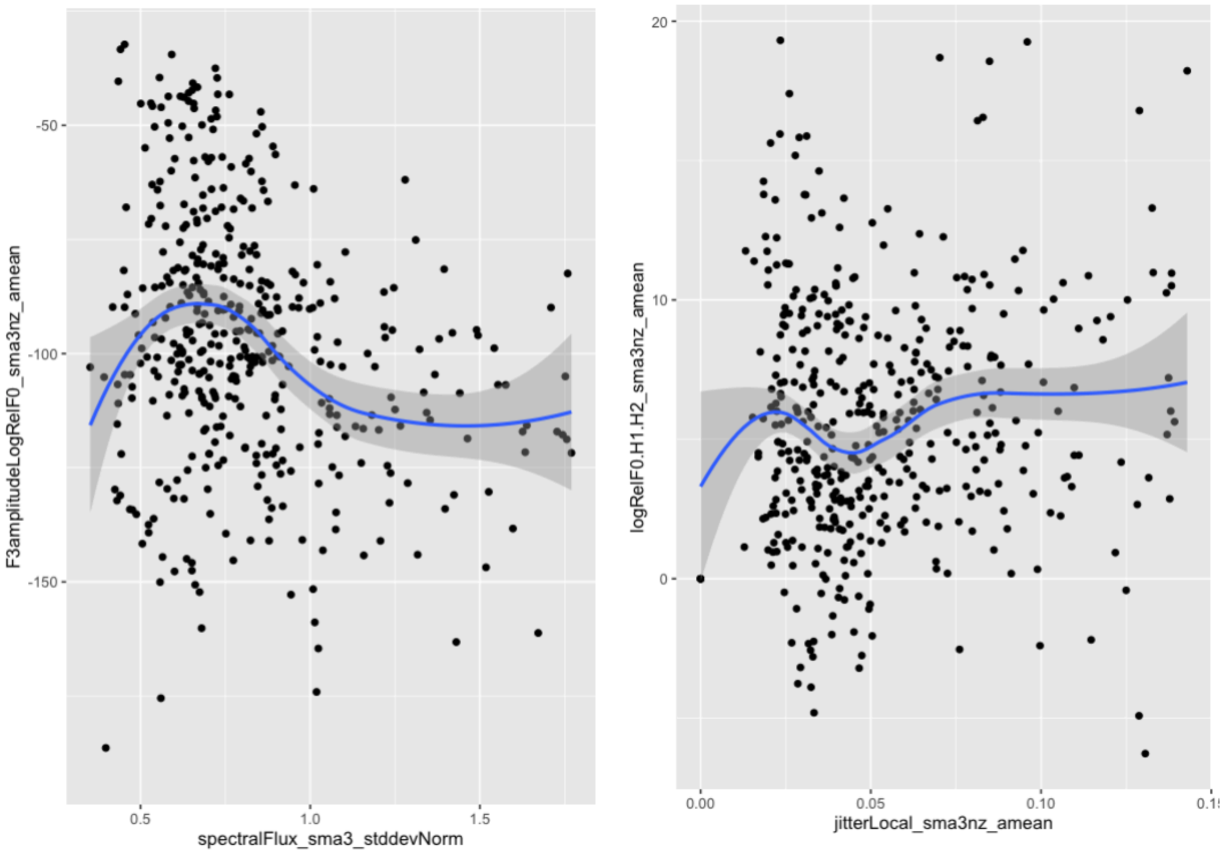


**Figure S2.** Non-monotonic associations between features.

### RESULTS

#### Performance using hyperparameter tuning

| | Features | LogisticRegression | MLP | RandomForest | SGDClassifier |
|---|---|---|---|---|---|
| Reading | 88 | .85 (.78–.93) | .87 (.8–.92) | .86 (.77–.91) | .86 (.75–.92) |
| Vowel | 88 | .86 (.77–.92) | .84 (.74–.92) | .84 (.77–.93) | .86 (.78–.91) |
| Reading+Vowel | 88 | .84 (.77–.90) | .86 (.78–.90) | .86 (.80–.92) | .83 (.76–.90) |

Table S1. Model performance after hyperparameter tuning increased performance by 0.01 on average across models and tasks.

#### Visualization of Redundant Features

See Figure S3–S11 for a visualization of redundant features for all participants, patients, and controls and for reading, vowel, and reading+vowel tasks. Top 5 features are highlighted in bold and their rank is displayed before the feature name with the corresponding leaf marked with an "x". When stratifying samples by disorder and task, clustering becomes more homogenous (clusters tend to contain a single feature type) in comparison to when all participants or both tasks are included as in Figure S5. Even in Figure S5, the chosen color-coded classification of features appears to be empirically replicated in this dataset given most low-level clusters (i.e., have higher dependency) are for the most part homogenous (i.e., of the same color). This also allows us to observe exceptions (e.g., mean spectral flux clusters with loudness features) which could otherwise be missed if using only a priori theoretical knowledge.

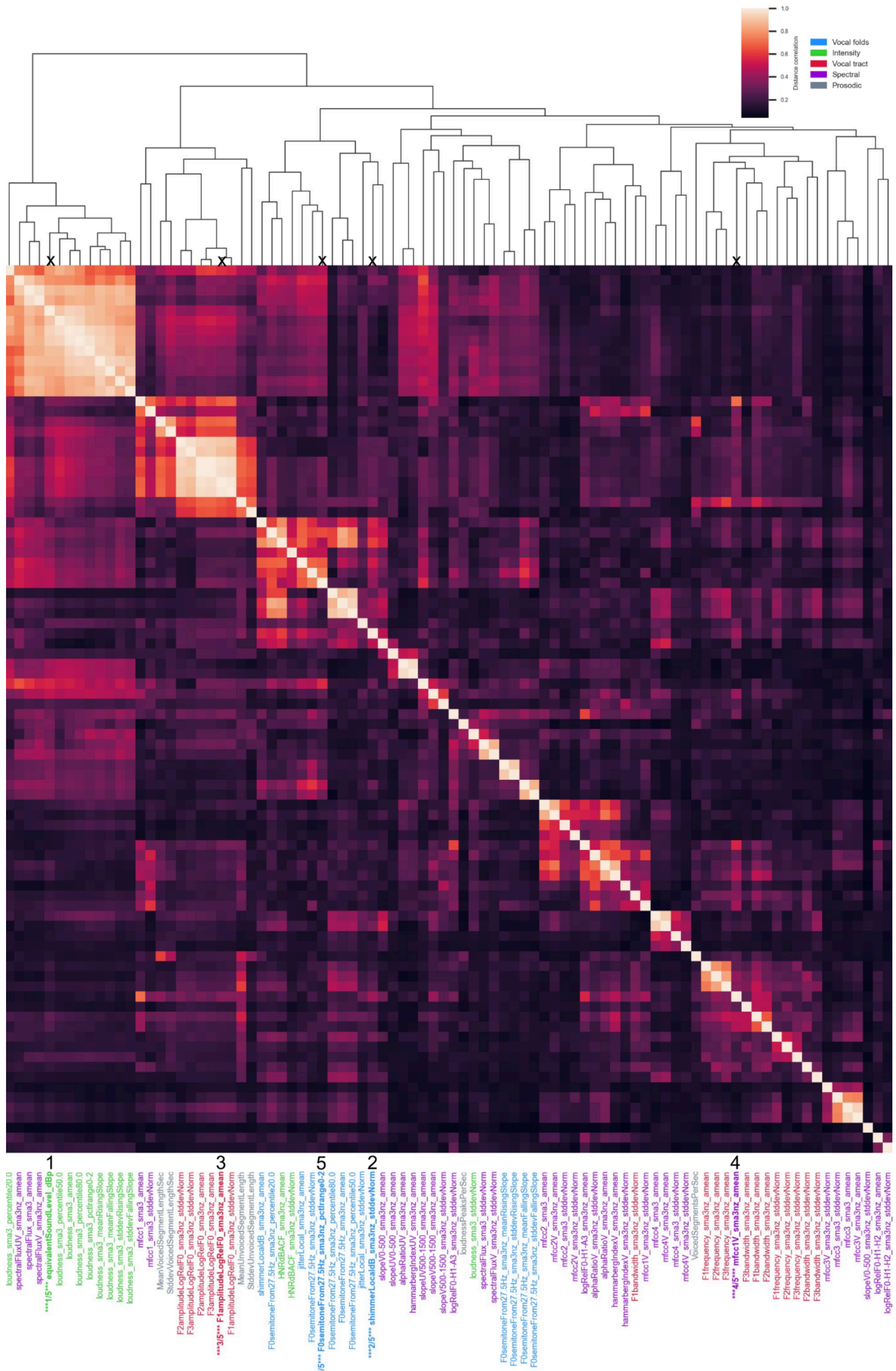


**Figure S3.** All participants, reading task: Visualization of features with shared information using pairwise distance correlation across the 88 eGeMAPs features. Squares are clusters of redundant features.

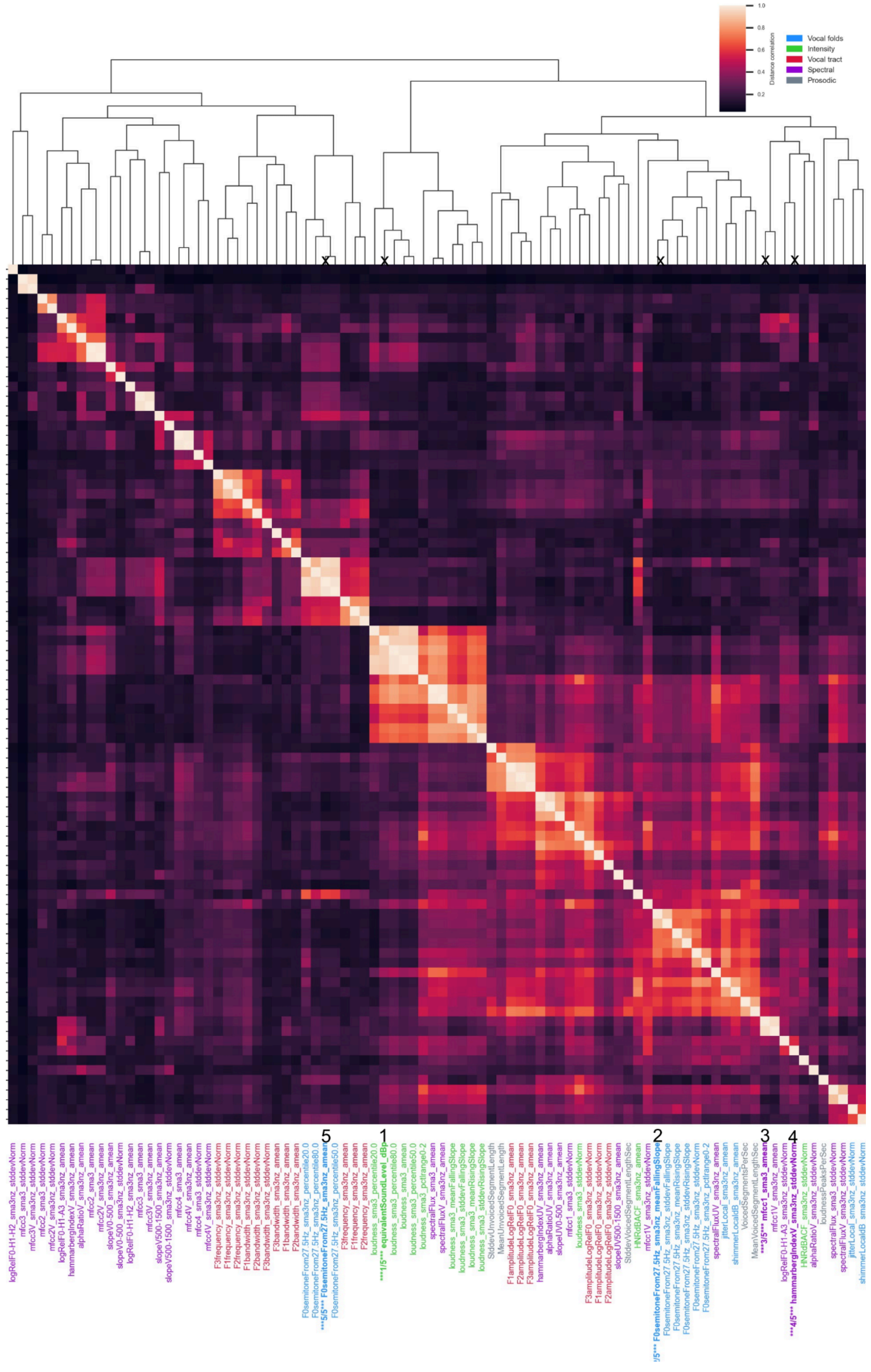


**Figure S4.** All participants, vowel task: Visualization of features with shared information using pairwise distance correlation across the 88 eGeMAPs features. Squares are clusters of redundant features.

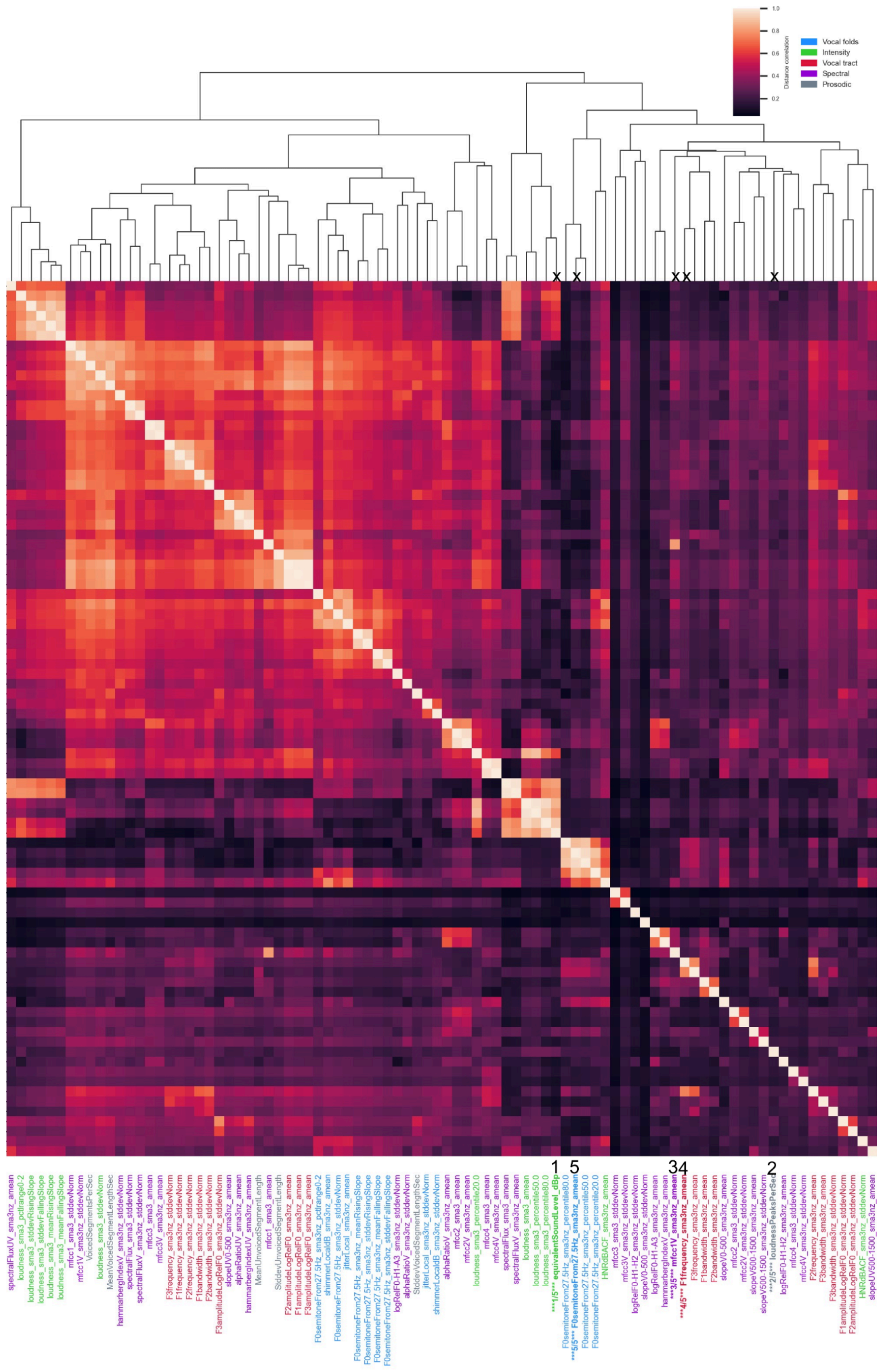

**Figure S5.** All participants, reading+vowel tasks: Visualization of features with shared information using pairwise distance correlation across the 88 features. Squares are clusters of redundant features.

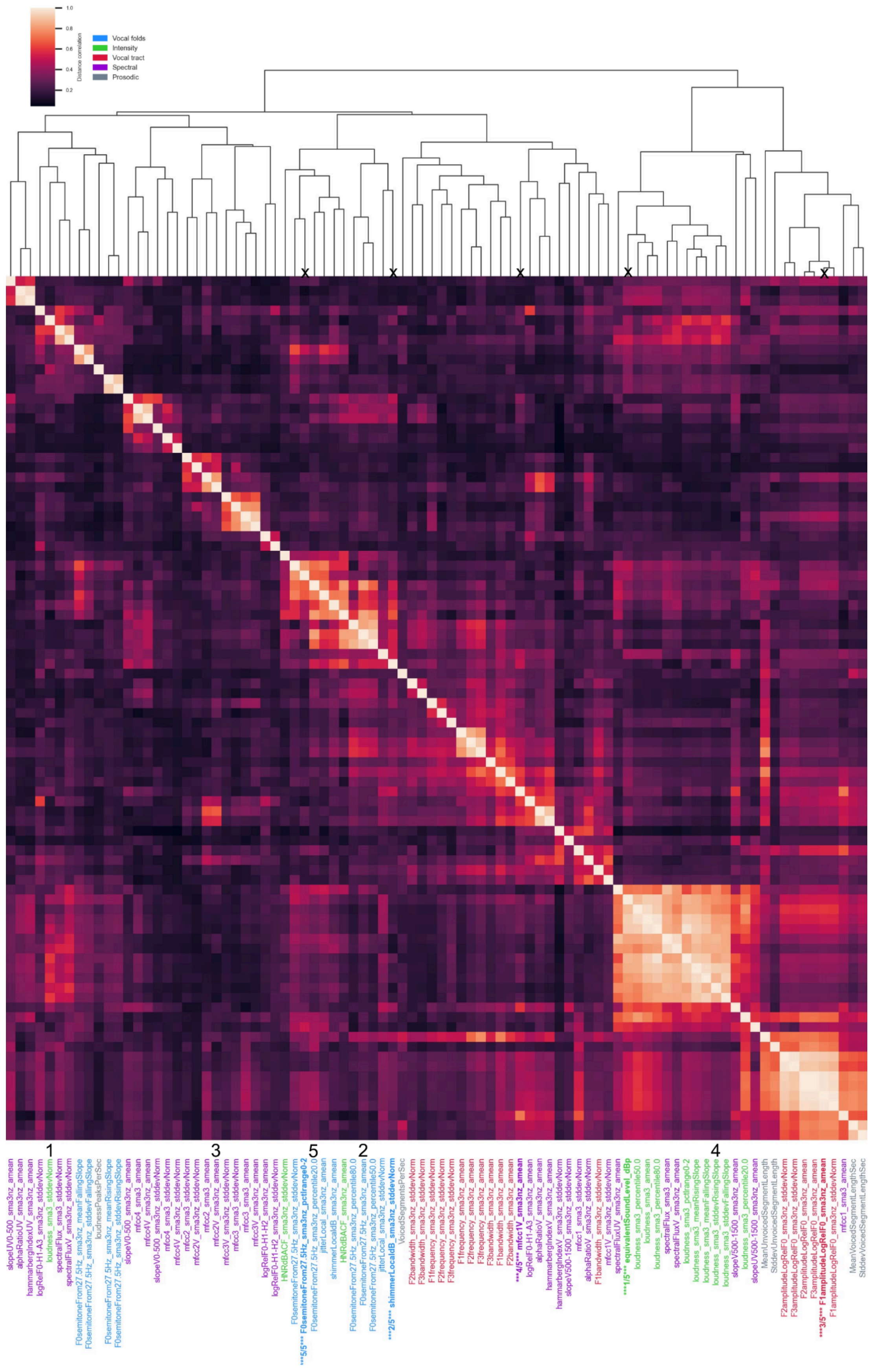


**Figure S6.** Patients, reading task: Visualization of features with shared information using pairwise distance correlation across the 88 eGeMAPs features. Squares are clusters of redundant features.

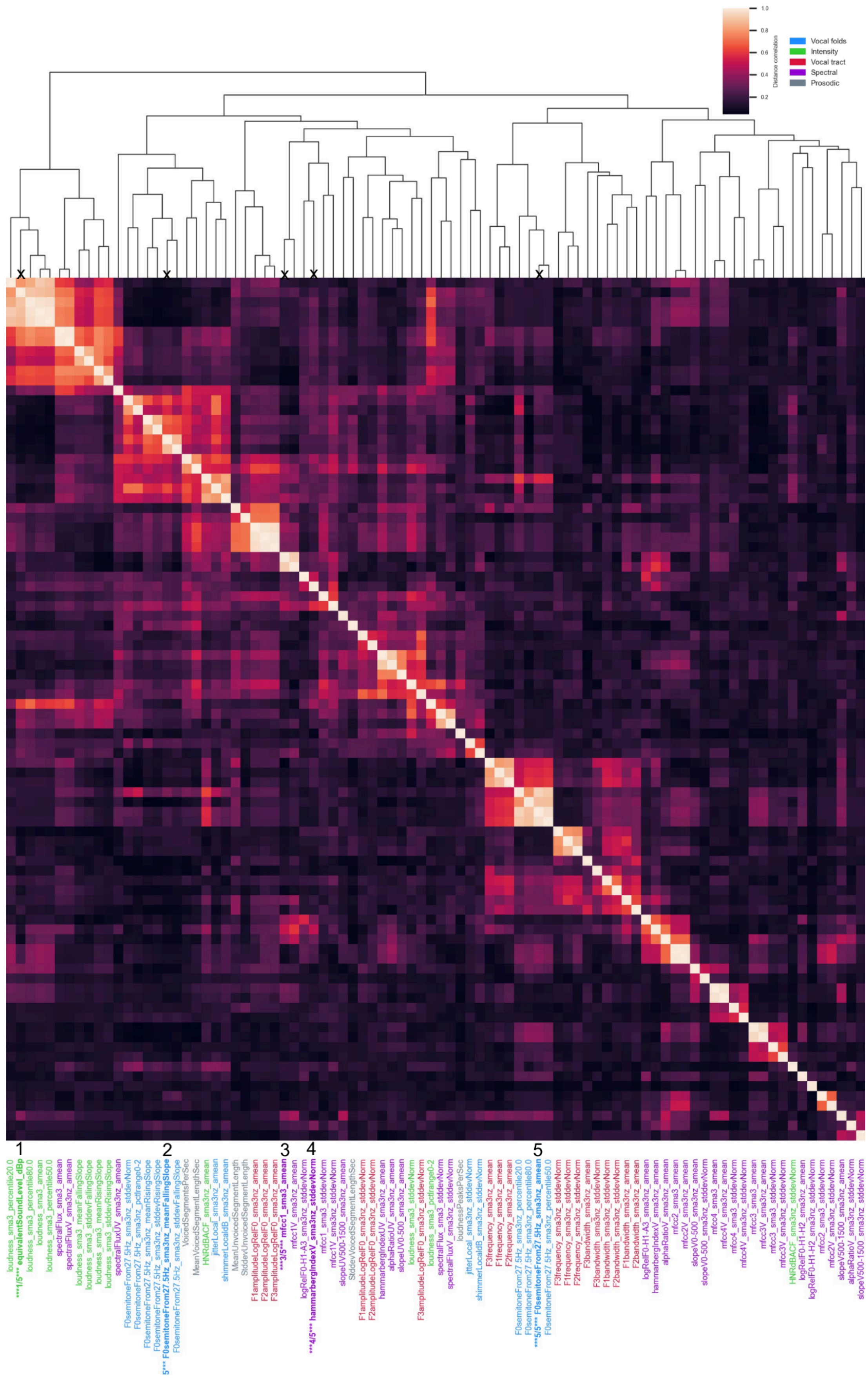

**Figure S7.** Patients, vowel task: Visualization of features with shared information using pairwise distance correlation across the 88 eGeMAPs features. Squares are clusters of redundant features.

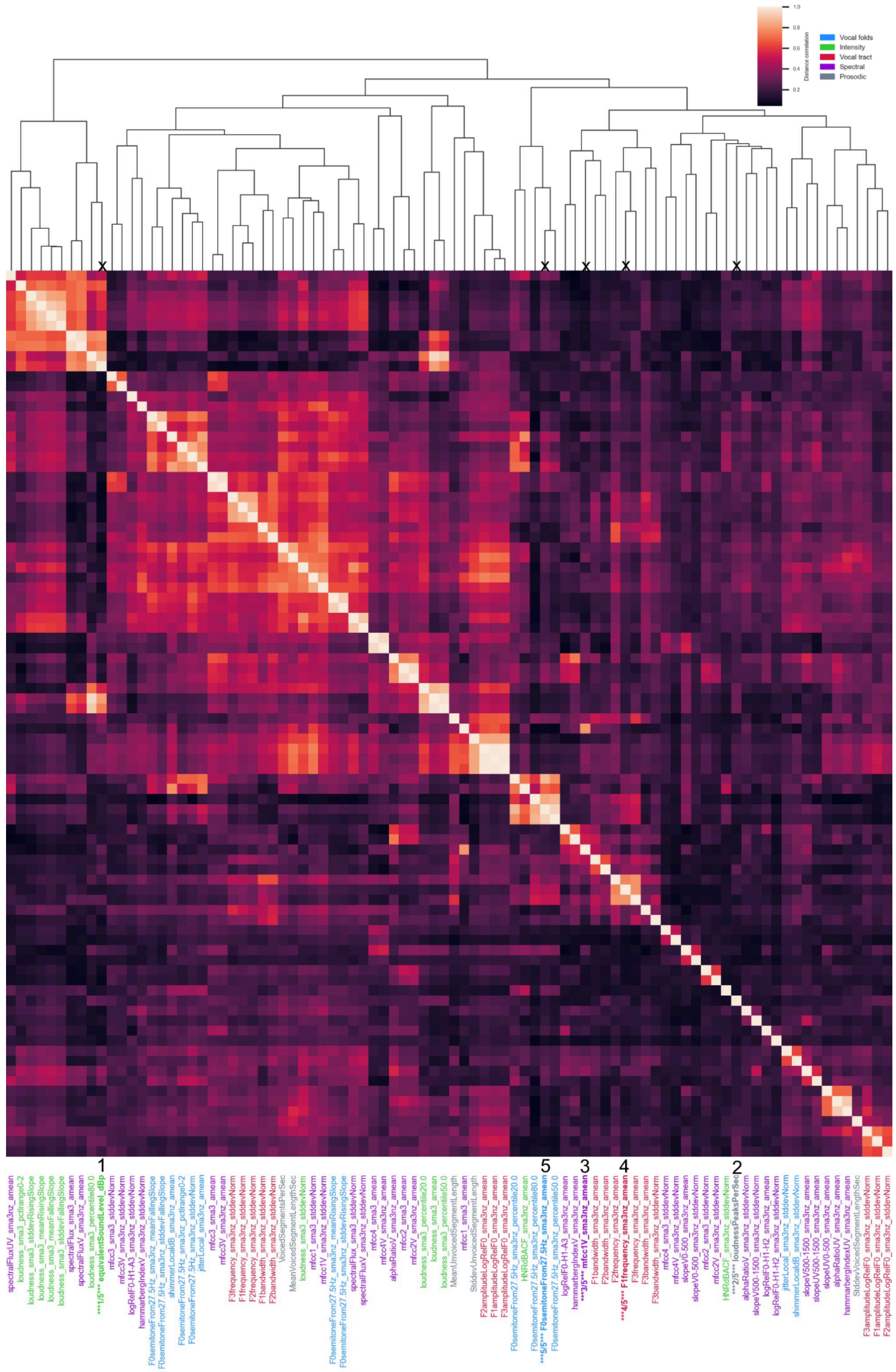


**Figure S8.** Patients, reading+vowel tasks: Visualization of features with shared information using pairwise distance correlation across the 88 eGeMAPs features. Squares are clusters of redundant features.

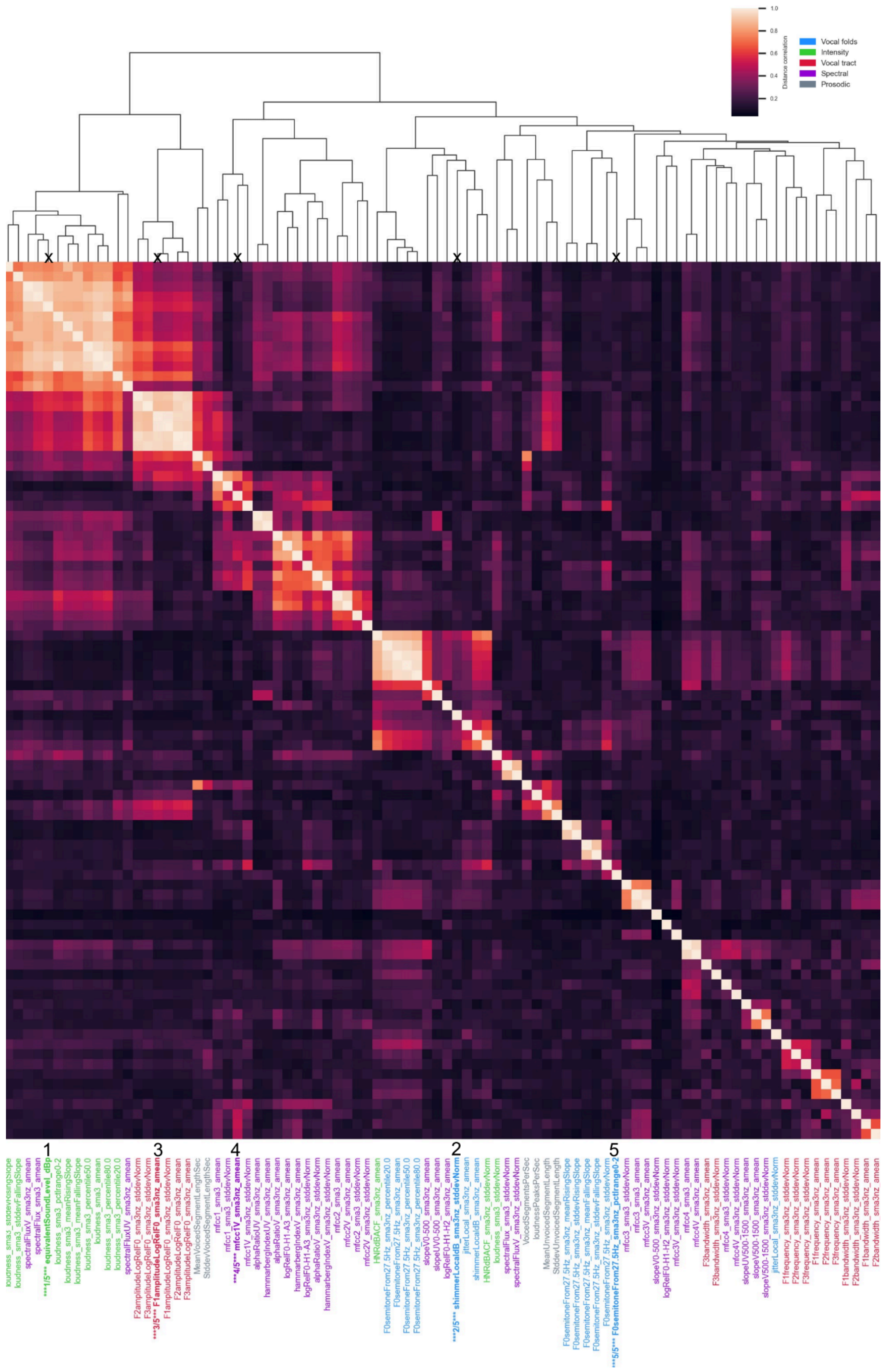


**Figure S9.** Controls, reading task: Visualization of features with shared information using pairwise distance correlation across the 88 eGeMAPs features. Squares are clusters of redundant features.

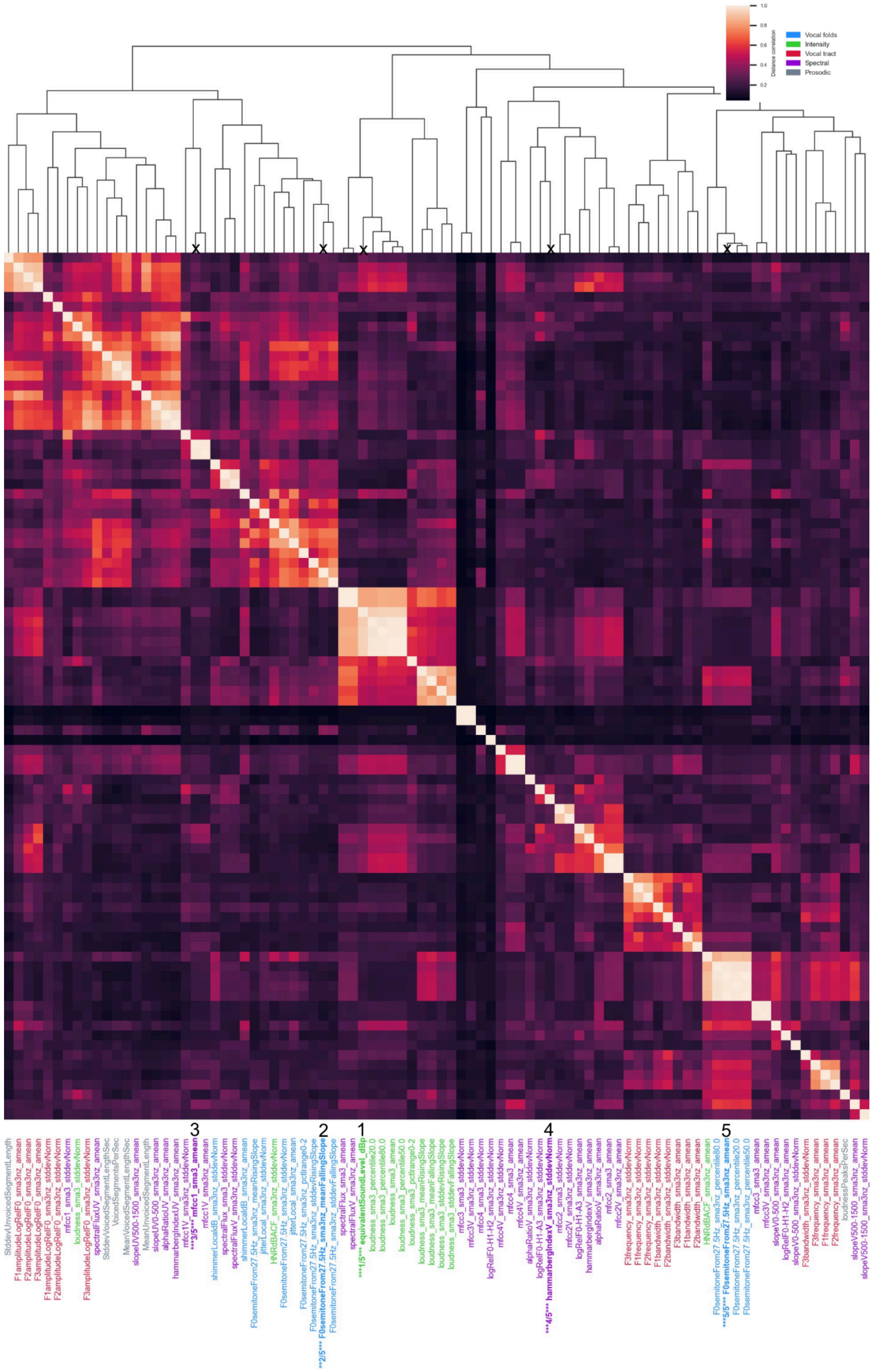


**Figure S10.** Controls, vowel task: Visualization of features with shared information using pairwise distance correlation across the 88 eGeMAPs features extracted. Squares are clusters of redundant features.

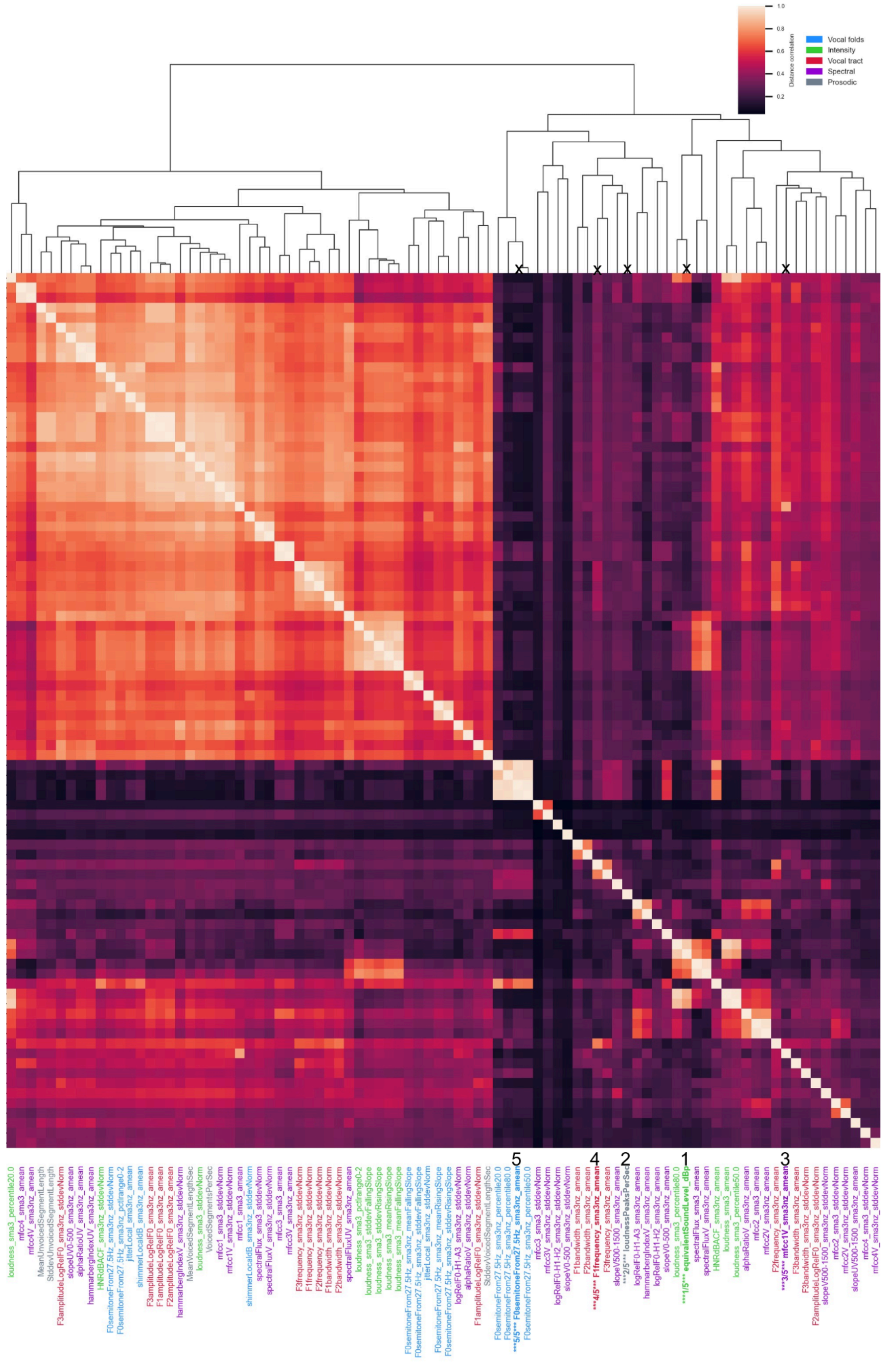

**Figure S11.** Controls, reading+vowel tasks: Visualization of features with shared information using pairwise distance correlation across the 88 features. Squares are clusters of redundant features.

#### Performance with and without redundant features

While removing redundant features is important for explainability, it should not be at the expense of predictive performance. Therefore, we trained and evaluated models and progressively removed redundant features to observe how performance dropped with fewer and fewer features. For each data type (reading, vowel, reading+vowel), through visual inspection of Figure S11, we chose the smaller feature set size that had similar performance to the full feature set size of 88 features: 39 features for reading, 13 features for vowel, and 19 features for reading+vowel.

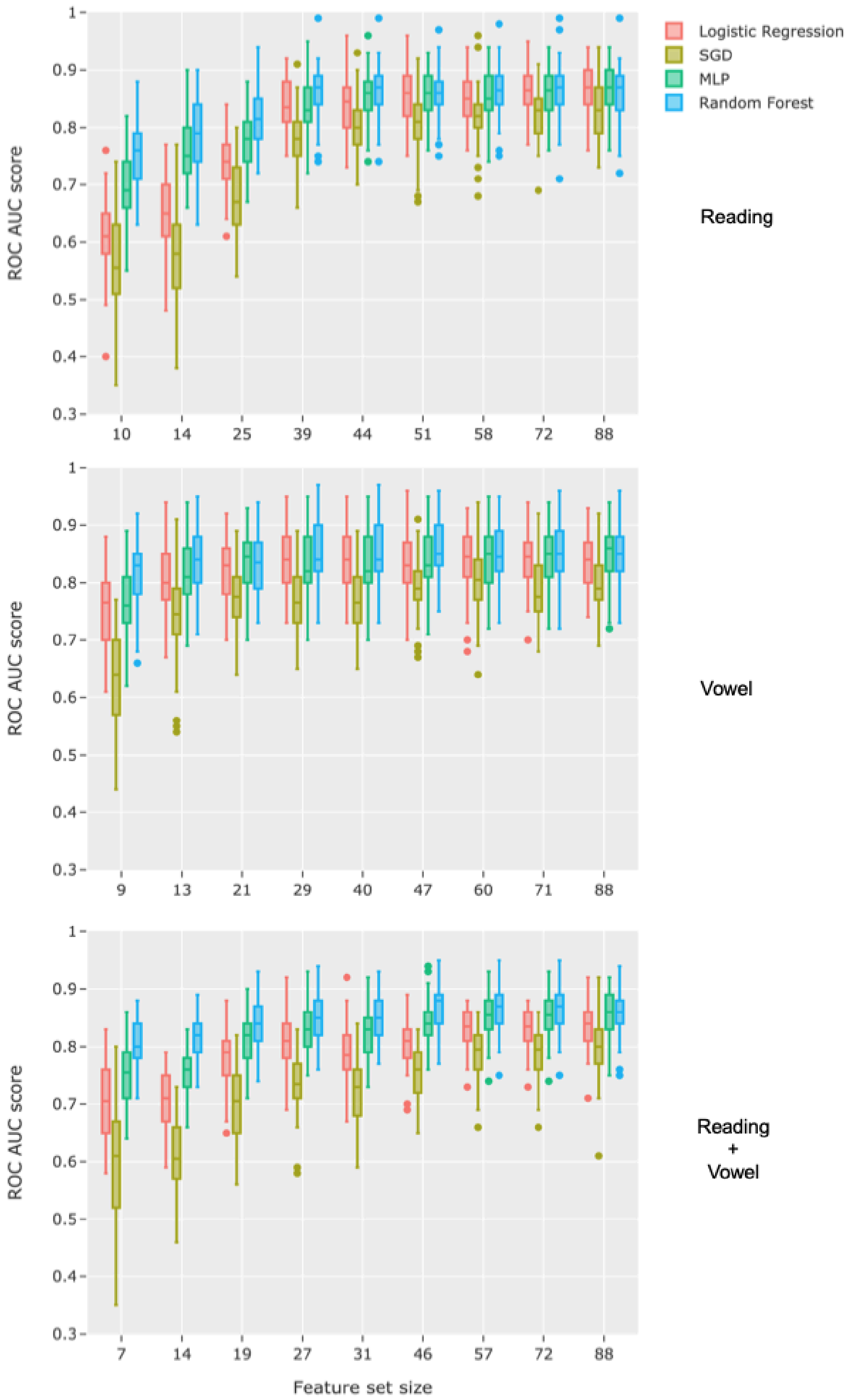


**Figure S12.** Performance as a function of feature set size using Independence Factor method for reducing feature redundancy. The feature sets remove features with distance correlation ≥ 0.2 up to 1.0 (i.e., keeping all features) in increments of 0.1.

### Feature Selection

To make sure information from the test sets is not having a strong influence on feature selection, we tested feature selection on 50 random train sets (80% of samples to match how models were trained) to make sure similar features were selected through this nested approach. If feature selection is relatively consistent across samples, removing features on the entire dataset should not be overfitting and is preferred for the explainability analysis to compare the same features. As seen in Table S2, all or most of the features used by selecting on the data set were also the most common across 50 splits and were selected in 91%, 83% and 76% of splits for reading, vowel and reading+vowel, respectively. Therefore, similar features are selected using both methods, but selecting on the entire dataset is preferred for explainability purposes (i.e., to rank the same features by their importance across all bootstrapping splits).

| Selection using | | Reading | Vowel | Reading+Vowel |
|---|---|---|---|---|
| Entire dataset | Optimal threshold and selected features | 0.5 | 0.3 | 0.4 |
| | Selected features | 39 | 13 | 19 |
| 50 bootstrap train sets | Selected features (mean [95% CI]) | 35.8 [34–38] | 12.3 [11–14] | 17.9 [16–20] |
| | Match between both methods (entire dataset / most common across 50 train sets) | 39/39 | 12/13 | 16/19 |
| | Selected in percentage of runs | 91% | 83% | 76% |

Table S2. Comparison of selecting features on the entire dataset (useful for explainability) versus selecting on 50 bootstrap (80–20) train splits. Original total features are 88. CI = Confidence Interval.

### Performance removing participants that used other recording system

Given 24 patients were recorded using an iPad, we trained models without their samples to make sure these differences in recordings were not driving performance. 66, 72, and 138 samples were removed from the reading, vowel, and reading+vowel datasets, respectively. Given the observed performance drop can also be due to removing training samples, the drop is not large enough to suspect that differences in recording are driving performance when using the full datasets (see Supplementary Table S3).

| | Features | LogisticRegression | MLP | RandomForest | SGD |
|---|---|---|---|---|---|
| Reading | 88 | .82 (.71–.87; .50) | .82 (.73–.88; .51) | .80 (.72–.88; .53) | .79 (.66–.87; .50) |
| Vowel | 88 | .78 (.71–.89; .50) | .79 (.68–.90; .54) | .81 (.73–.90; .52) | .74 (.60–.85; .45) |
| Reading+Vowel | 88 | .79 (.70–.87; .50) | .81 (.74–.88; .52) | .81 (.73–.88; .52) | .77 (.67–.84; .50) |

Table S3. Performance of models without 24 patients recorded on iPad. Median ROC AUC score from 50 bootstrapping splits (90% confidence interval; median score of null model). The control group represents 60% of the training samples. MLP: Multi-Layer Perceptron; SGD: Stochastic Gradient Descent Classifier.

Furthermore, we tested how well a model trained on all participants except those using the iPad and tested on the 24 UVFP patients that used the iPad (see Table S4) to assess generalizability of the model to different recording settings. However, since the iPad recordings were all patients we can therefore only measure false negative rate but not ROC AUC. We used only the controls matched in age and sex to the remaining UVFP patients for training the models to maintain a balanced dataset (i.e., 53 UVFP patients and 53 matched controls).

| | Features | LogisticRegression | MLP | RandomForest | SGD |
|---|---|---|---|---|---|
| Reading | 88 | 0.08 | 0.26 | 0.09 | 0.39 |
| Vowel | 88 | 0.1 | 0.11 | 0.36 | 0.12 |
| Reading+Vowel | 88 | 0.12 | 0.2 | 0.12 | 0.12 |

Table S4. False negative rate (FNR) of training on one recording device and testing on 24 UVFP patients that used iPad. FNR is generally quite low. Performance can also be influenced by having a smaller training set in order to balance the classes.

#### Biased features

We identified features that are biased (differ between groups not due to the intrinsic nature of UVFP, equivalentSoundLevel_dBp, and the other intensity-related features it is strongly associated with: loudness_sma3_amean, loudness_sma3_stddevNorm, loudness_sma3_percentile20.0, loudness_sma3_percentile50.0, loudness_sma3_percentile80.0, loudness_sma3_pctlrange0-2, loudness_sma3_meanRisingSlope, loudness_sma3_stddevRisingSlope, loudness_sma3_meanFallingSlope, loudness_sma3_stddevFallingSlope, loudnessPeaksPerSec, equivalentSoundLevel_dBp, HNRdBACF_sma3nz_amean, and HNRdBACF_sma3nz_stddevNorm.

We correlated these intensity features with all other features and removed the 43 features that had a distance correlation > 0.3 as seen in Table S5. We correlated the audio duration with all other features and removed the 44 features that had a distance correlation > 0.3 as seen in Table S5.

| **Features associated with intensity features** | **dcor** |
|---|---|
| spectralFluxV_sma3nz_amean | 0.9 |
| spectralFlux_sma3_amean | 0.89 |
| F0semitoneFrom27.5Hz_sma3nz_amean | 0.88 |
| F0semitoneFrom27.5Hz_sma3nz_percentile20.0 | 0.88 |
| F0semitoneFrom27.5Hz_sma3nz_percentile50.0 | 0.86 |
| F0semitoneFrom27.5Hz_sma3nz_percentile80.0 | 0.82 |
| spectralFluxUV_sma3nz_amean | 0.78 |
| slopeUV500-1500_sma3nz_amean | 0.7 |
| F0semitoneFrom27.5Hz_sma3nz_stddevNorm | 0.57 |

| | |
|---|---|
| slopeV500-1500_sma3nz_amean | 0.57 |
| spectralFlux_sma3_stddevNorm | 0.54 |
| shimmerLocaldB_sma3nz_amean | 0.53 |
| shimmerLocaldB_sma3nz_stddevNorm | 0.48 |
| slopeV0-500_sma3nz_amean | 0.47 |
| F3frequency_sma3nz_amean | 0.46 |
| VoicedSegmentsPerSec | 0.45 |
| F0semitoneFrom27.5Hz_sma3nz_pctlrange0-2 | 0.44 |
| F1bandwidth_sma3nz_amean | 0.43 |
| F2frequency_sma3nz_amean | 0.43 |
| MeanUnvoicedSegmentLength | 0.42 |
| F2amplitudeLogRelF0_sma3nz_amean | 0.4 |
| F3amplitudeLogRelF0_sma3nz_amean | 0.4 |
| jitterLocal_sma3nz_amean | 0.39 |
| F2amplitudeLogRelF0_sma3nz_stddevNorm | 0.39 |
| F3amplitudeLogRelF0_sma3nz_stddevNorm | 0.39 |
| F1frequency_sma3nz_amean | 0.38 |
| jitterLocal_sma3nz_stddevNorm | 0.38 |
| MeanVoicedSegmentLengthSec | 0.37 |
| F1frequency_sma3nz_stddevNorm | 0.37 |
| mfcc1_sma3_amean | 0.37 |
| F1amplitudeLogRelF0_sma3nz_amean | 0.36 |
| spectralFluxV_sma3nz_stddevNorm | 0.35 |
| F1amplitudeLogRelF0_sma3nz_stddevNorm | 0.35 |
| F2frequency_sma3nz_stddevNorm | 0.35 |
| StddevUnvoicedSegmentLength | 0.34 |
| mfcc4_sma3_amean | 0.34 |
| alphaRatioV_sma3nz_stddevNorm | 0.33 |
| mfcc4V_sma3nz_amean | 0.33 |

| | |
|---|---|
| hammarbergIndexV_sma3nz_stddevNorm | 0.32 |
| F0semitoneFrom27.5Hz_sma3nz_meanFallingSlope | 0.32 |
| F3bandwidth_sma3nz_stddevNorm | 0.31 |
| F3frequency_sma3nz_stddevNorm | 0.31 |
| F2bandwidth_sma3nz_amean | 0.31 |
| mfcc2_sma3_amean | 0.3 |

**Table S5. Features with distance correlation (dcor) > 0.3 with biased intensity-related features.**

In Table S6, we report the distance correlations between all 88 features with audio duration to observe which of the features is associated with the observed systematic bias between groups (Figure 2). Among the highest associations, we find the longer the recording the higher the intensity/loudness variables, which is expected given all control recordings are 3.5 s while the only recordings above this duration are all UVFP. Our clinician ratings confirmed that UVFP have higher background noise and perceived louder speech due to recording biases that we've discussed. Therefore, being a UVFP patient is likely confounding the association between duration and loudness (likely causing both to be higher and therefore to be associated). Therefore, all distance correlations with audio duration could be spurious or partially biased because they can also be explained by differences between UVFP and controls. Therefore, we do not think these distance correlations with audio duration should be overinterpreted for potential ways duration could be affecting features beyond UVFP-control differences. Of course, after trimming audio duration so that all samples are the same length, there is no variation in duration to compute these correlations; this specific bias is mitigated.

| **Features associated with audio duration** | **dcor** |
|---|---|
| equivalentSoundLevel_dBp | 0.76 |
| spectralFlux_sma3_amean | 0.72 |
| loudness_sma3_stddevRisingSlope | 0.71 |
| loudness_sma3_amean | 0.7 |
| spectralFluxV_sma3nz_amean | 0.7 |
| spectralFluxUV_sma3nz_amean | 0.7 |
| loudness_sma3_percentile80.0 | 0.69 |
| slopeUV500-1500_sma3nz_amean | 0.67 |
| loudness_sma3_percentile50.0 | 0.67 |

| | |
|---|---|
| loudness_sma3_meanRisingSlope | 0.65 |
| loudness_sma3_percentile20.0 | 0.65 |
| loudness_sma3_pctlrange0-2 | 0.64 |
| loudness_sma3_stddevFallingSlope | 0.64 |
| loudness_sma3_meanFallingSlope | 0.6 |
| F0semitoneFrom27.5Hz_sma3nz_pctlrange0-2 | 0.54 |
| F0semitoneFrom27.5Hz_sma3nz_stddevNorm | 0.53 |
| jitterLocal_sma3nz_amean | 0.5 |
| slopeV500-1500_sma3nz_amean | 0.46 |
| loudnessPeaksPerSec | 0.46 |
| F0semitoneFrom27.5Hz_sma3nz_percentile20.0 | 0.45 |
| hammarbergIndexUV_sma3nz_amean | 0.44 |
| alphaRatioUV_sma3nz_amean | 0.43 |
| HNRdBACF_sma3nz_stddevNorm | 0.42 |
| F0semitoneFrom27.5Hz_sma3nz_meanFallingSlope | 0.42 |
| logRelF0-H1-A3_sma3nz_stddevNorm | 0.42 |
| HNRdBACF_sma3nz_amean | 0.41 |
| F0semitoneFrom27.5Hz_sma3nz_stddevRisingSlope | 0.4 |
| spectralFluxV_sma3nz_stddevNorm | 0.39 |
| F0semitoneFrom27.5Hz_sma3nz_stddevFallingSlope | 0.37 |
| F0semitoneFrom27.5Hz_sma3nz_meanRisingSlope | 0.36 |
| spectralFlux_sma3_stddevNorm | 0.34 |
| shimmerLocaldB_sma3nz_amean | 0.33 |
| StddevVoicedSegmentLengthSec | 0.32 |
| shimmerLocaldB_sma3nz_stddevNorm | 0.31 |

**Table S6. Features with distance correlation (dcor) > 0.3 with biased audio duration.**
